# Resolving the classification rates and molecular architecture of early-onset chronic kidney disease with NephVar

**DOI:** 10.64898/2026.07.22.26358680

**Authors:** Joshua P. Pillai, John A. Sayer, the NephVar Renal Registry

## Abstract

The genetic architecture of early-onset chronic kidney disease (CKD) is caused by more than 200 monogenic genes, where their common diagnostic classes include congenital anomalies of the kidney and urinary tract, steroid-resistant nephrotic syndrome, nephronophthisis-related ciliopathies, chronic glomerulonephritis, and urinary stone disease. While advancements in whole-exome and whole-genome sequencing have enabled identification of disease-causing variants, their rates of classification have remained unknown. Likewise, the molecular effects of these pathogenic variants remain unresolved, which is essential for improving personalized treatment approaches. In this study, we collected clinical and biophysical data from 117,373 genetic variants across 129 monogenic genes causing early-onset CKD. This data established the NephVar registry, which aims to be a molecular dictionary for nephrologists to classify variants and resolve their unique molecular effects. Through NephVar, we estimated 1-15% of alleles are reclassified and the time to reclassification per variant is 2-12 years in early-onset CKD. Furthermore, NephVar identified the molecular effects of all variant types, emphasizing missense variants. Our analyses indicate that intrinsically disordered regions of proteins are protective against disease-causing missense alleles across most diagnostic classes, but often occur through a buried loss-of-function (LoF) mechanism. Additionally, we show that the mode of inheritance for these monogenic genes influences clustering patterns of genetic variants, where autosomal dominant (AD) genes are more clustered than those of autosomal recessive (AR) genes. This data accurately predicted the non-LoF effects in *INF2*, *PAX2*, *GATA3*, *ACTN4*, and *LMX1B* causing inherited nephrotic syndromes. We demonstrate that variant effect prediction is effective for downgrading variants of unknown significance and classifying AR genes, but challenging for pathogenic alleles in AD genes. Lastly, we propose standards and guidelines for determining non-LoF effects, including gain-of-function and dominant negative, in inherited nephrotic syndrome. Overall, the NephVar renal registry has important implications for defining the molecular architecture and estimating the progress of molecular diagnostics for early-onset CKD.

## Introduction

Early-onset chronic kidney disease (CKD) is defined as an estimated glomerular filtration rate of less than 60 mL/min/1.73 m^2^ for at least 3 months or the presence of kidney damage up to 25 years of age, where its progression to kidney failure is managed with kidney transplantation or dialysis (Vivante and Hildebrandt., 2016). From huge experimental efforts using whole-exome and whole-genome sequencing strategies in the last two decades, over 200 monogenic genes have been linked to early-onset CKD, which accounts for an estimated 70% of cases (Vivante and Hildebrandt., 2016). From these genes, 130 have been established in next generation sequencing (NGS) genetic screening panels for inherited kidney disease (Bullich et al., 2018). Typically, these gene panels are used to screen congenital anomalies of the kidney and urinary tract (CAKUT), steroid-resistant nephrotic syndrome (SRNS), nephronophthisis-related ciliopathies (NPHP), chronic glomerulonephritis (CGN), and urinary stone disease (USD) (Rodriguez., 2014; Vats et al., 2013; Walawender et al., 2023). Overall, there has been exceptional progress towards diagnostic efforts for early-onset CKD, but the genetic architecture of this early-onset kidney phenotype remains incompletely understood. Resolution of these etiologies are essential to guiding the development of gene therapies and other precision therapeutics to prevent progressive CKD (Peek and Wilson., 2023).

However, these diagnostic efforts are limited by two key factors: (1) a lack of functional studies delineating the molecular effects of identified genetic / genomic variants and (2) small sample sizes in comparative genetic and mechanistic studies. For instance, biallelic missense alleles in *CD2AP*, encoding the glomerular protein CD2-associated protein, have been reported in SRNS, while heterozygous loss-of-function (LoF) variants appear to act as susceptibility alleles or modifiers in focal segmental glomerulosclerosis (FSGS). In addition, several single nucleotide polymorphisms risk variants in *CD2AP* have been robustly associated with Alzheimer’s disease in genome-wide association studies (Tao et al., 2019; Benoit et al., 2010). As of June 2026, no pathogenic missense variants in *CD2AP* have been reported in ClinVar under the American College of Medical Genetics and Genomics/Association for Molecular Pathology (ACMG/AMP) guidelines (Richards et al., 2015). Instead, almost all reported missense alleles are classified as variants of uncertain significance (VUSs), creating a major obstacle for comparative studies because well-defined pathogenic and benign control groups are lacking. Consequently, for *CD2AP* missense alleles, and many other monogenic kidney disease genes, the etiological basis of clinical phenotypes remains poorly understood, limiting their potential for clinical translation.

Furthermore, the prevalence and classification rates of reported genetic variants in monogenic disease-causing genes remain largely uncharacterized. These estimates are essential for guiding public policy, informing investment in genomic infrastructure, and prioritizing therapeutic translational efforts globally over the coming decades. The broader goal is to advance the field toward achieving ‘precision nephrology’, in which the mechanistic and pathway-level effects of genetic variants are systematically resolved, analogous to large-scale historical efforts observed in precision oncology to functionally characterize somatic alleles across all oncogenes and tumor suppressor genes (Muiños et al., 2021; Wang., 2016).

Lastly, there are technical barriers that prevent geneticists, nephrologists, and clinicians from obtaining molecular-level resolution of genetic variants in these monogenic kidney genes. In particular, methods in structural biology could reveal how variants broadly alter the wild-type protein structures, localization, or function, but remain underutilized in clinical investigations. Resolving these genotype patterns may help reveal the underlying mechanisms for genotype-phenotype correlations in early-onset CKD.

In this work, we developed NephVar, a renal registry for clinical and biophysical data of genetic variants from 129 monogenic genes causing early-onset CKD. NephVar helped estimate the classification and reclassification rates per ACMG/AMP guidelines. We also used NephVar to reveal broad disease-causing mechanisms of missense variants. Finally, this study provides molecular clues for resolving VUSs and predicting their pathogenicity at scale. Together, these findings establish NephVar as the first unified framework enabling nephrologists and geneticists to systematically determine the molecular effects of genetic variants in monogenic kidney disease.

## Results

### The NephVar renal registry provides a molecular dictionary to help classify and characterize genetic variants in early-onset chronic kidney disease

In this study, we introduce the NephVar renal registry (nephvar.github.io/NephVar) on behalf of the NephVar Registry. This renal registry is a molecular dictionary for use by clinical nephrologists and geneticists for the classification of inherited kidney diseases. The NephVar platform is a unified system for known monogenic kidney disease genes that on a single gene level allows the user to (1) define the classification history of a genetic variant in a known kidney disease gene and (2) establish its predicted molecular effects. Currently, NephVar has 117,373 unique genetic variant data for 129 monogenic genes causing early-onset CKD. For completeness, we included genomic deletions and duplications for each diagnostic class—CAKUT, CGN, NPHP, SRNS, and USD— culminating in 118,367 alleles in the data registry. For each variant, NephVar provides data for the ClinVar submission histories, laboratories, phenotypes, pathogenicity, and reclassification histories of reported alleles **(Figure S1-2)**. The platform also provides biophysical data, especially useful for missense alleles, including the burial state of altered amino acid residues, domain annotation, secondary structural elements, and the grammars of intrinsically disordered regions (IDRs) of these predicted proteins.

In **Figure 1A**, we have provided a schematic design of NephVar, showing an overview of the systems and methods integrated from the NCBI ClinVar, AlphaFold, UniProt, and NARDINI+ databases. Given the dynamic nature of variant submission histories, we plan to update individual gene data biannually and expand the system for additional monogenic kidney gene panels (e.g. cystic kidney disease, renal tubulopathies) in future versions. This update policy is appropriate as the frequency of submissions per variant is 1-2 submissions across each diagnostic class **(Figure 1B)**.

**Figure 1.**
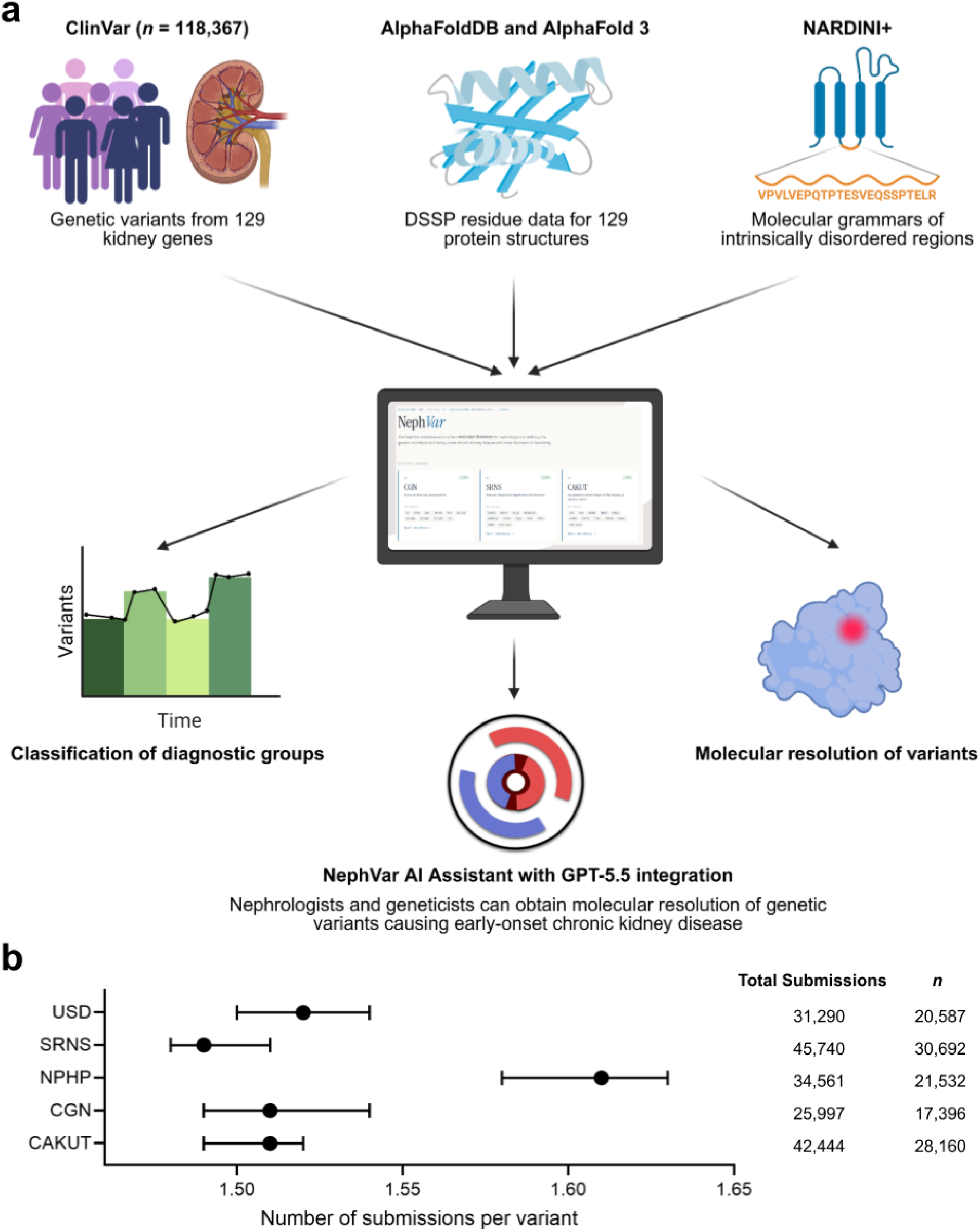
Design of the NephVar registry and submission rates for early-onset chronic kidney disease. **(a)** The NephVar registry sources its clinical and biophysical data from ClinVar, AlphaFoldDB, and the NARDINI+ algorithm. These datasets are then analyzed and reported in the NephVar database (https://nephvar.github.io/NephVar/) that provides molecular resolution of genetic variants and classification rates for the 5 diagnostic groups. Additionally, nephrologists and geneticists may use the NephVar AI Assistant powered by GPT-5.5 for analysis and reasoning of the variant data. **(b)** The submission rates per genetic variant and submission frequencies for each diagnostic class. All data have been presented as the mean and 95% CI. The illustration in (a) has been created using BioRender at www.biorender.com. USD: Urinary stone disease; SRNS: steroid-resistant nephrotic syndrome; CAKUT: congenital anomalies of the kidney and urinary tract; NPHP: nephronophthisis-related ciliopathies; CGN: chronic glomerulonephritis; NARDINI: Non-random Arrangement of Residues in Disordered Regions Inferred using Numerical Intermixing.

Furthermore, to allow for direct usage among non-technical users, we integrated the NephVar data registry with GPT-5.5 to create the ‘NephVar AI Assistant’, where nephrologists and geneticists may obtain detailed molecular resolution of genetic variants of interest. As a blind validation test, we prompted the AI Assistant to help resolve variants identified from a cohort of Omani patients with nephrotic syndrome who underwent whole-exome sequencing with nephrotic syndrome **(Figure S3-4)** (Al Riyami et al., 2026). Using prompt engineering methods, we designed AI prompts shown in **Figure S3-4**. Preliminary studies with GPT-5.5 demonstrate model reasoning using both NephVar and the available resources via the internet in conjunction to efficiently define a variant of interest. However, in **Figure S4**, we show that the model can be susceptible to inaccuracies in scanning through the data, thereby suggesting its usage as a secondary resource to the NephVar registry itself. We plan to update the training parameters of the NephVar AI Assistant biannually similar to the registry.

### NephVar estimates the classification rates of genetic variants in all diagnostic groups causing early-onset chronic kidney disease

After establishing NephVar, we were interested in estimating the classification rates and time to reclassification for genetic variants from each diagnostic group (*n* = 118,367). For genetic variants with at least a single submission, we define a reclassification as a change in ACMG classification between the first and latest submission within ClinVar. For instance, *CD2AP* E525del was first classified as LP, but its latest resubmission was downgraded to VUS, and thereby considered an ACMG reclassification. For alleles in the CAKUT diagnostic group, 2.4% (95% CI: 2.21-2.57) were ever reclassified (*n* = 662). In **Figure 2A**, we estimated reclassification rates for CAKUT alleles from their initial ACMG classification, where VUSs were the lowest class ever reclassified within 4-9 years. For CGN, the reclassification rate was estimated at 4.17% (95% CI: 3.88-4.48) (*n* = 710). The times to reclassification spanned from 2-6 years, where VUSs had the lowest rates **(Figure 2B)**. NPHP reclassification was also similar with 3.21% (95% CI: 2.98-3.46) (*n* = 686) with LB/B and VUSs remaining the lowest rates **(Figure 2C)**. This estimated rate remains consistent for SRNS with a reclassification rate of 3.23% (95% CI: 3.03-3.43) (*n* = 979), ranging from 2-9 years and VUSs having the lowest rates **(Figure 2D)**. Lastly, the rates for USD were closest to CGN at 4.12% (95% CI: 3.85-4.40) from 4-10 years **(Figure 2E)**. Overall, from all diagnostic groups in **Figure 2**, VUS and LB were the highest classified alleles, respectively. These times and rates to reclassification corroborate the reclassification rates identified in a cohort study of 3.2 million patients by Labcorp Genetics (Kobayashi et al., 2024).

**Figure 2.**
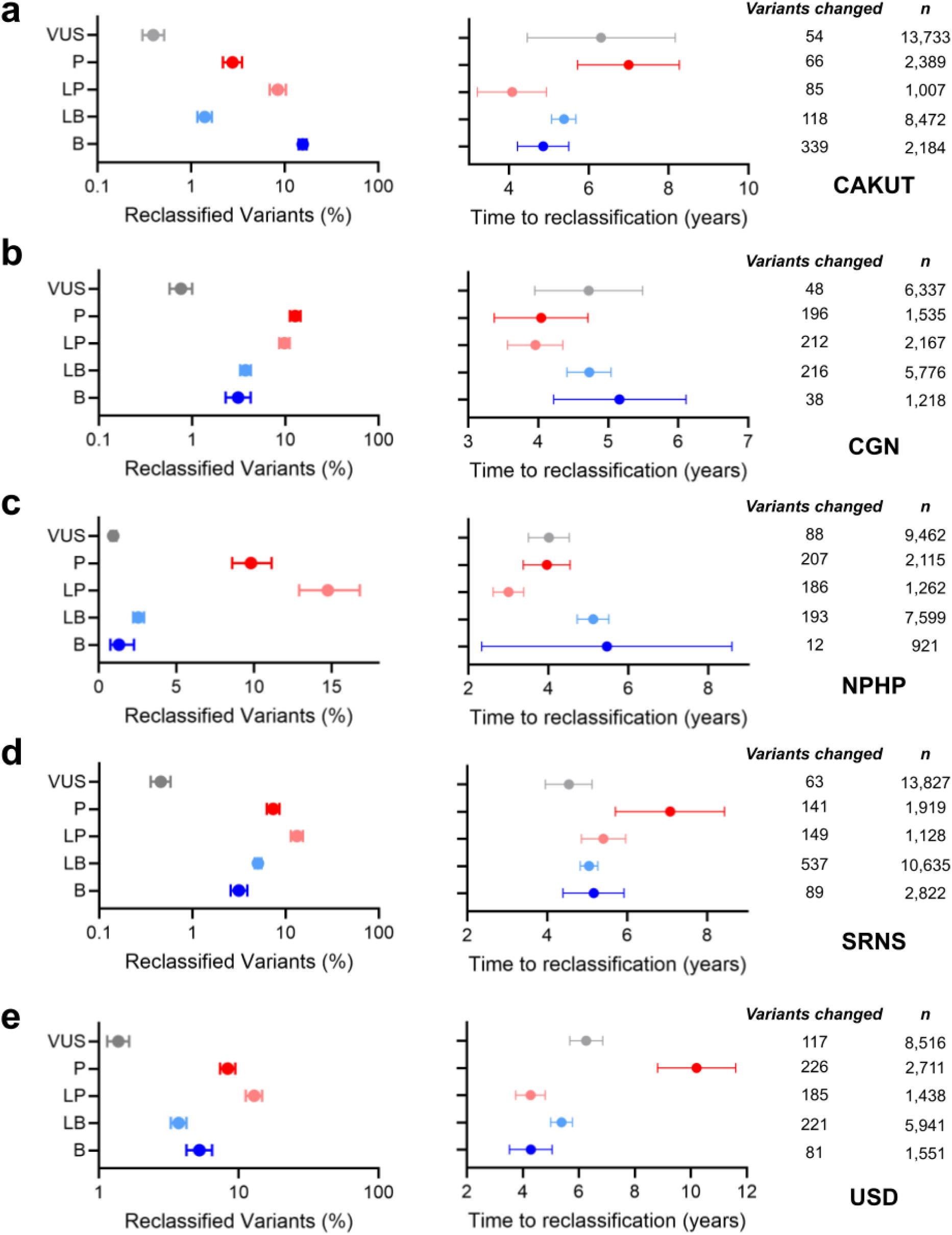
Reclassification rates and time to reclassification of diagnostic classes in early-onset CKD. **(a)** Rates for genetic variants in CAKUT, where VUSs remain the lowest group to be reclassified and the time to reclassification ranges from 2-8 years. **(b)** Rates for CGN, where VUSs remain the lowest reclassified group and the time to reclassification is from 4-6 years. **(c)** Rates for NPHP, where both benign and VUSs are the lowest classified groups. Most classes had a time to reclassification of 4 years with exception to benign and likely benign at 5 years. **(d)** Rates for SRNS, where VUSs are the lowest, similar to CGN and CAKUT. **(e)** Lastly, rates for USD, where VUSs are the lowest reclassified group, but time to reclassification remains the longest at 10 years for pathogenic variants. All data have been presented as the mean and 95% CI, along with their sample sizes (*n*). P: pathogenic; LP: likely pathogenic; VUS: variant of unknown significance; LB: likely benign; B: benign.

From all unique genetic variants (*n* = 117,373), we identified 25,497 alleles with at least 2 submissions, of which 3,637 alleles differed in their initial and latest classification. In **Table S1**, we provide the directions of reclassification for each class. For LP/P alleles, the strength of clinical evidence increases and genetic variants are usually upgraded. For VUSs, alleles are mostly downgraded, though most remain unresolved. Lastly, for LB/B alleles, their classifications are usually downgraded.

### Missense variants positioned in intrinsically disordered regions are protective against disease-causing effects except chronic glomerulonephritis

Next, we were interested in defining the association of missense variants in each diagnostic kidney disease group with their position in intrinsically disordered regions (IDR) of the predicted proteins. Missense variants in IDRs are particularly challenging to assess biophysically due to their lack of structural data and often discordant predictions among variant effect predictors (Fawzy and Marsh., 2025). In **Figure 3A**, we illustrate our study design assessing whether ACMG classifications differ based on the position of a missense variant in or out of an IDR. From 129 genes, NephVar identified 74 with IDRs. There were 32,064 missense variants identified that were further stratified by position inside or outside IDRs. Before comparative studies, we hypothesized that missense variants in IDRs had a lower frequency of LP/P classifications compared to non-IDR regions due to their minimal effects on the stickers-and-spacers framework (Martin et al., 2020).

**Figure 3.**
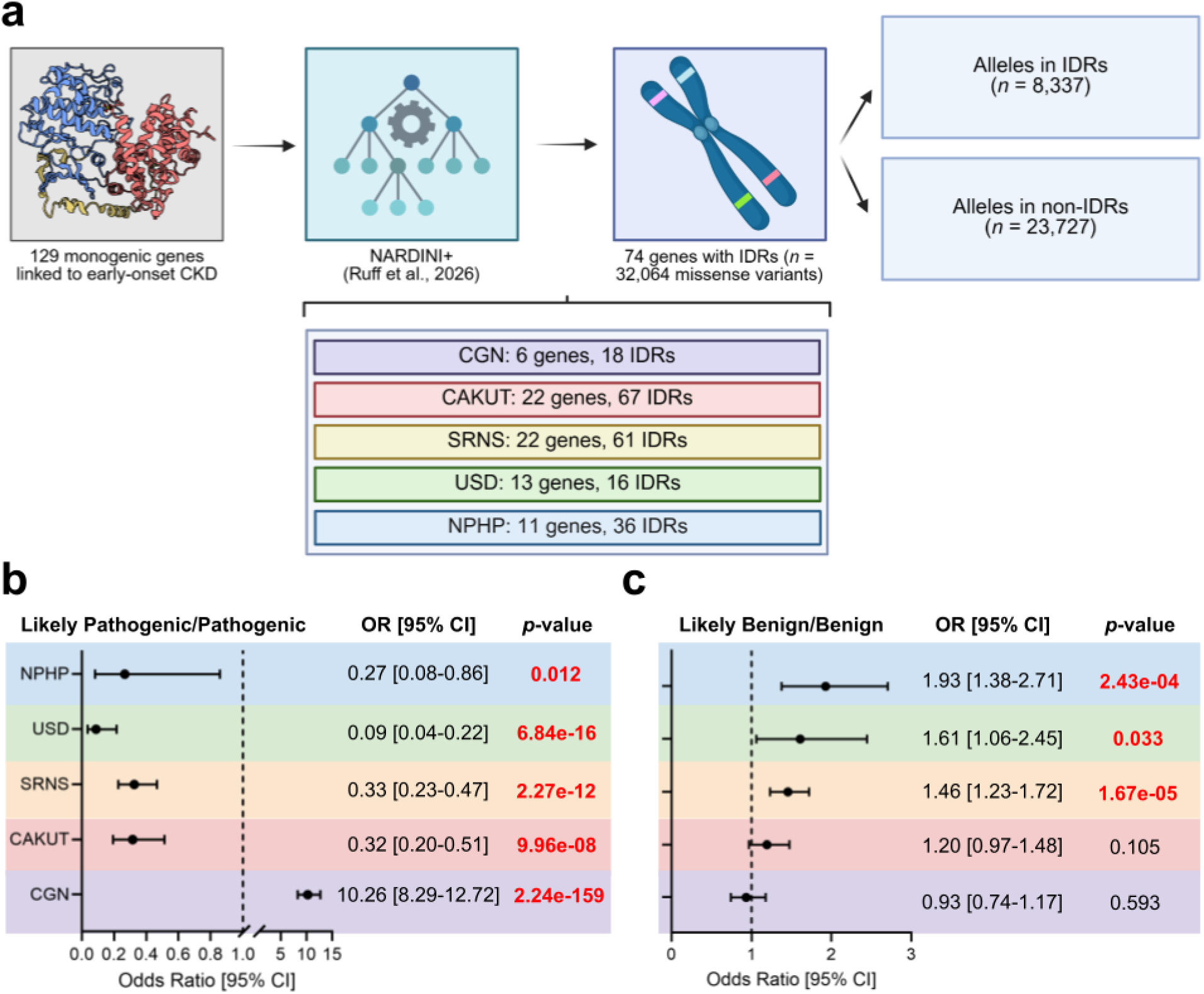
Enrichment analysis of missense variants in intrinsically disordered regions (IDRs). **(a)** Schematic design for the enrichment analyses. From all 129 monogenic genes, we obtained the IDR regions available for 74 genes. We then identified 32,064 missense variants occurring in these 74 genes, and stratified into alleles found within the IDR or outside the IDR. The gene and IDR frequencies for each diagnostic class is also provided. **(b)** Enrichment analysis of LP/P missense variants in IDRs shows all diagnostic groups are protective against these variants except for CGN. **(c)** Enrichment analysis of LB/B missense variants in IDRs, which is non-significant in CAKUT and CGN, but significantly higher odds in SRNS, USD, and NPHP. All *P* values are indicated in the corresponding figure. All data have been presented as the mean and 95% CI. The illustration in (a) was created using BioRender at www.biorender.com.

The statistical framework of this study involved calculating the frequencies of LB/B and LP/P missense variants and the odds ratio and variance using the Woolf logit method followed by Fisher’s exact test for significance. We show the statistical model of LP/P alleles from IDR regions in **Figure 3B**, where IDRs in all diagnostics groups but CGN had a 67-91% lower odds of a pathogenic missense allele affecting the region. Similarly, we show the LB/B model **(Figure 3C)** that had a 46-93% higher odds of benign alleles occurring in IDRs for SRNS, USD, and NPHP. Most importantly, there is a strong odds of pathogenic variants occurring in IDRs of CGN while benign remains insignificant with non-IDRs, respectively. The key drivers of this strong association are variants in the *COL4A5* gene causing X-linked Alport syndrome. In NephVar, the collagen alpha-5 (IV) chain contains a single IDR that spans 85.20% of the protein, which contains 60% (*n* = 672) of LP/P alleles. Pathogenic missense variants in the collagen alpha-3 (IV) chain and collagen alpha-4 (IV) chain encoded by *COL4A3* and *COL4A4* were also drivers with 38% (*n* = 327) and 31% (*n* = 273) of alleles found in IDRs but not as strongly associated as *COL4A5* variants. It is well established that the IDRs of type IV collagen molecules are essential for network assembly, cross-linking, and the triple helical domain for the G-X-Y motifs, so the mechanism for this strong association of CGN is resolved (Fidler et al., 2018; Sundaramoorthy et al., 2002).

### Pathogenic missense variants associated with early-onset CKD are often buried and impose loss-of-function effects except chronic glomerulonephritis

Given the low pathogenicity of genetic variants observed in IDRs, we were particularly interested in resolving the broad molecular effects of all remaining missense variants. Indeed, there are no postulates in regards to the disease-causing mechanisms of these variants in causing early-onset CKD. From all monogenic genes, we identified 50,425 missense alleles (including the variants in IDRs) aligning with the canonical isoforms shown in NephVar, where 87.29% were classified as VUSs (*n* = 44,021). We hypothesized that LP/P missense variants imposed a LoF mechanism affecting protein folding, and therefore calculated the relative solvent accessibility (RSA) of all residue positions for the variant cohort. In **Figure 4A**, the solvent exposure of benign missense variants was 0.47 (95% CI: 0.44-0.49) that was significantly higher than pathogenic at 0.33 (95% CI: 0.31-0.35) (*p* = 0.0045). Both are above the threshold of burial (RSA = 0.25), but understandably due to combined diagnostic groups.

**Figure 4.**
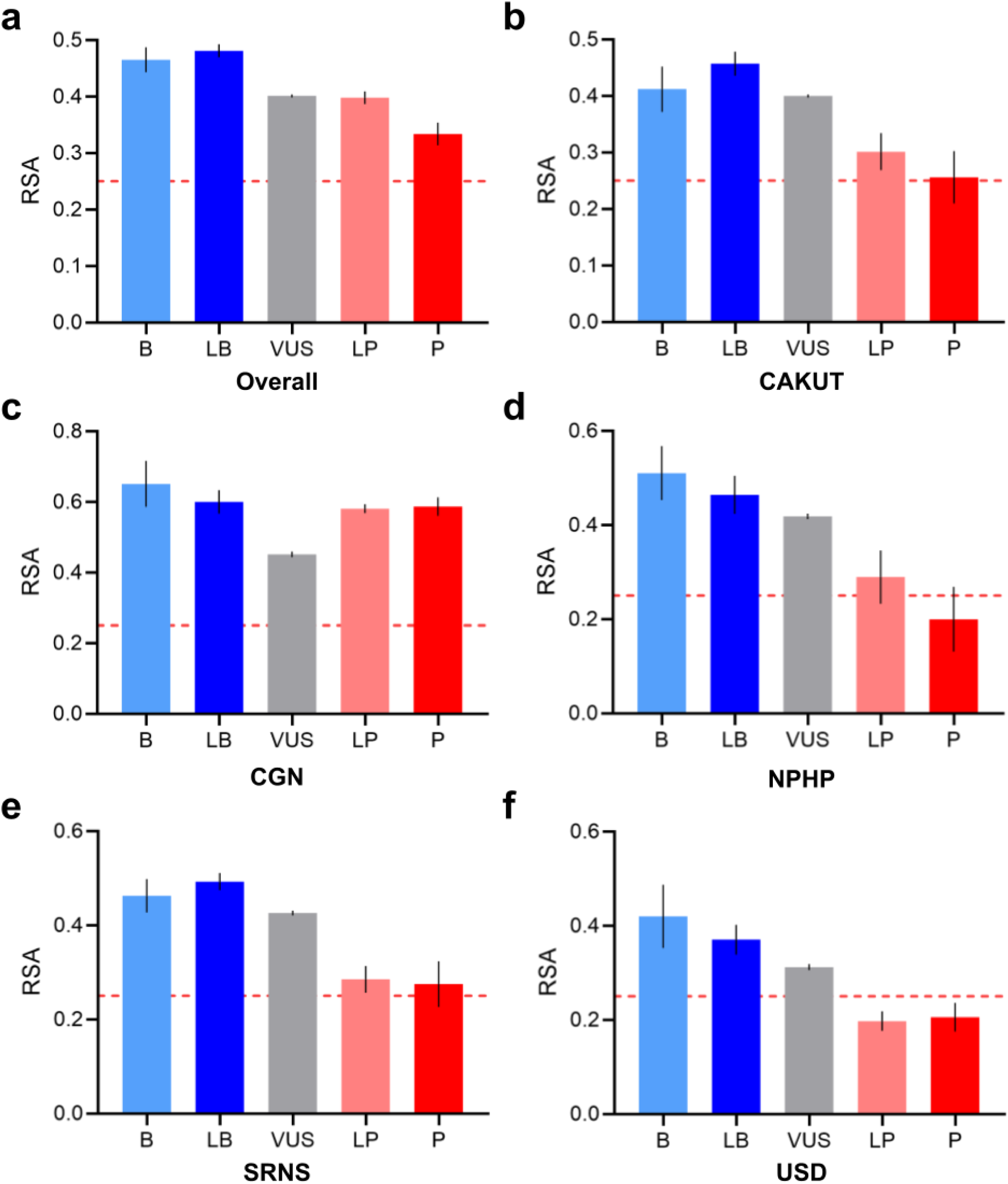
Relative solvent accessibility (RSA) of missense variants in all diagnostic classes for early-onset CKD. **(a)** The overall RSA of all ACMG classes are all surface-exposed. **(b)** In CAKUT, missense variants in each functional class are exposed with exception to the pathogenic (P) class. **(c)** The correlation observed in (a) for the surface-exposure in LP/P alleles can be resolved from the CGN group with highly exposed LP/P variants. **(d)** Missense variants in NPHP follow a similar pattern to all groups but CGN, where LP/P are at or below the burial threshold. Again, these trends are consistent in both **(e)** SRNS and **(f)** USD. All data have been presented as the mean and 95% CI. The dotted red line indicates the burial threshold for RSA.

In **Figure 4B-F**, we stratified the missense variants by diagnostic classes. In all groups except CGN, LP and P variant subsets were non-significant from the burial threshold. Of course, the high surface-exposure in CGN is owing to *COL4A3/4/5* alleles that had the highest reported missense variants of all monogenic genes. Therefore, we observe a strong association of LP/P in buried residue positions, thereby suggesting a LoF effect affecting protein folding. Furthermore, we investigated the structural elements each mutational class affected **(Table S2)**. Pathogenic alleles in CGN strongly affected loops at 92-94%, CAKUT spread across all three, SRNS and USD with helices and loops, and NPHP spread across all types. Our correlations suggest, similar to RSA, mutational patterns in the affected secondary structure are clustered for disease-causing variants. From all LP/P missense variants excluding those in CGN, 61.70% (*n* = 1,239) were buried in their respective protein structures **(Table S3)**. However, what exactly are the molecular mechanisms underlying these surface-exposed pathogenic variants? Using the domain annotations and established protein-protein interfaces, we resolved 45.6% of all surface pathogenic alleles in **Table S4**. Overall, buried LoF mechanisms are dominant, but those affecting the surface interfaces are incompletely understood.

We are also aware of the valid criticism of AlphaFold predictions as *in silico* models in NephVar, but the ‘60-40’ ratio of buried to surface pathogenic missense alleles is likely consistent for individual genes with some variation. *NPHS1* encoding nephrin is the perfect gene candidate due to its balanced variant cohort, where we observed a 67.9-32.1 split in the resolution studies of its fishnet architecture in the slit diaphragm (Pillai and Sayer., 2026; Birtasu et al., 2025). In **Figure S5**, predicted Local Distance Difference Test (pLDDT) scores have been provided for all genes, a metric indicative of the confidence of each structural prediction, where most are accurate. Furthermore, our analyses of all remaining variant types, including synonymous, intronic, and frameshift alleles, were unremarkable for their predicted disease-causing mechanisms from established literature, and their estimated frequencies are in **Table S5-7**.

### Pathogenic variants in monogenic genes associated with autosomal dominant patterns of inheritance are highly clustered compared those of autosomal recessive in early-onset CKD

After resolving the molecular effects for most variant types, we were interested in defining the role of all modes of inheritance (MOI) with their disease-causing effects in early-onset CKD. Using a modified extent of disease clustering (EDC) metric from Gerasimavicius et al., (2022), we quantified the effect of the MOI stratified by LP/P and LB/B alleles. ‘EDC’ is a simple quantitative metric to assess clustering of variants of interest per gene, detailed in the **Methods and Materials**.

In **Figure 5A**, we assessed the clustering of LB/B variants across all MOIs, where we show expected non-significance between autosomal dominant (AD) and recessive (AR) patterns (*p* = 0.7884). We also show the individual EDC scores for all monogenic genes with at least 3 LB/B variants (including X-linked, AD/AR, etc), where most are randomly scattered throughout the protein structures (EDC ∼ 1) **(Figure 5B)**. The rare instances of scores below 1 observed in *COL4A5* (X-linked) and *INF2* are not ‘anti-clustering’, but due to alleles spanning the entire sequence. Conversely, in **Figure 5C**, we detected significantly higher clustering of LP/P alleles in monogenic genes following AD inheritance compared to AR. Again, we show individual EDC scores per monogenic gene, where LP/P alleles of *INF2*, *LMX1B*, and *GATA3* following AD inheritance are highly clustered **(Figure 5D)**. Accounting for match pairing, we also identified that LP/P alleles are more often clustered than LB/B in both AD (*p* = 0.0001; *n* = 20) and AR (*p* = 0.0106; *n* = 48). Therefore, we have strong evidence to suggest that alleles in monogenic genes causing early-onset CKD are often clustered when following the AD MOI compared to AR. Due to the sample size of X-linked and AD/AR MOIs, we did not include these genes for individual statistical testing.

**Figure 5.**
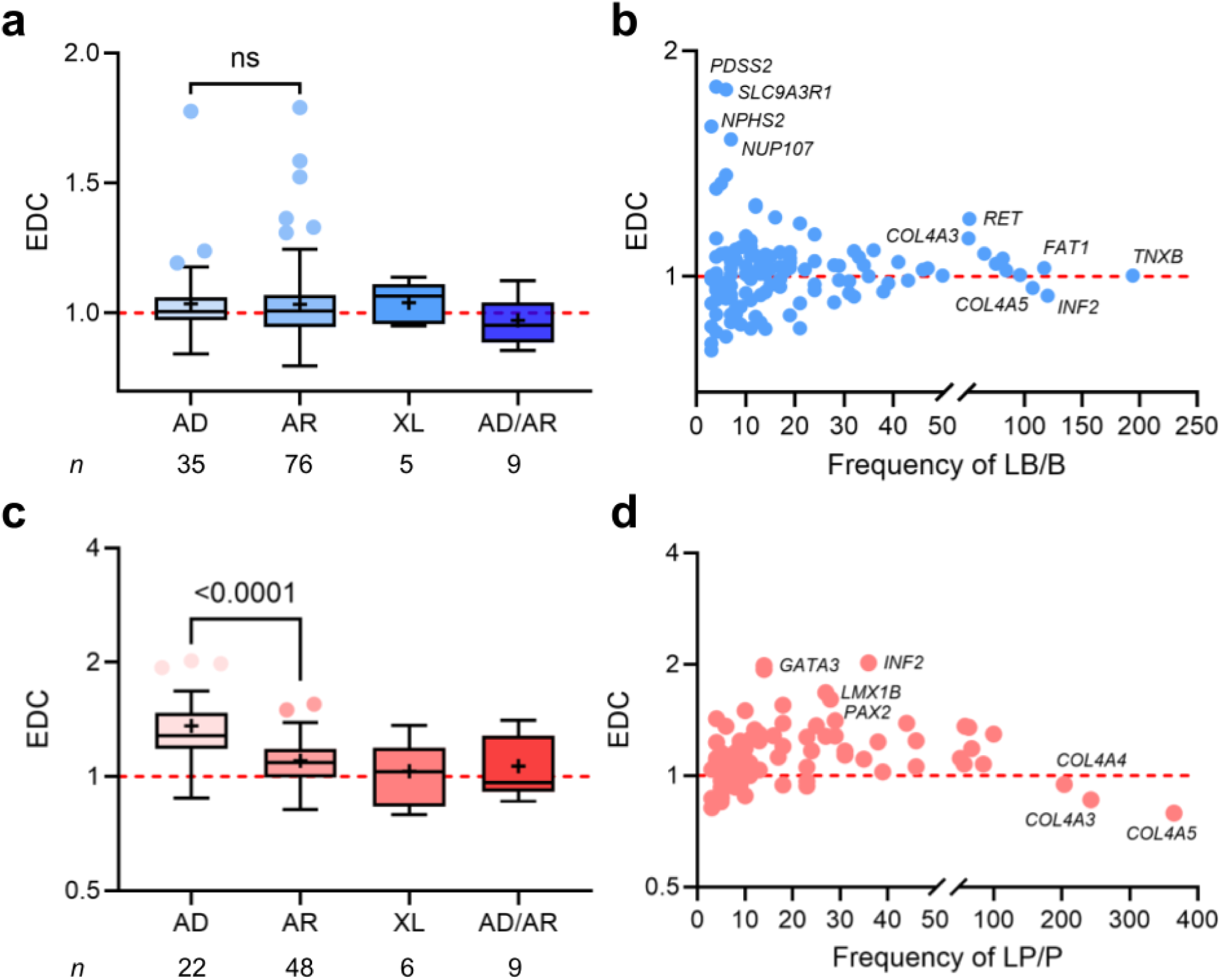
The extent of disease clustering (EDC) stratified by mode of inheritance in early-onset CKD. **(a)** EDC is non-significant for LB/B missense alleles in both autosomal dominant (AD) and autosomal recessive (AR). **(b)** The EDC per monogenic gene with their LB/B frequencies are generally close to 1, indicating random clustering for all monogenic genes. **(c)** Likewise, EDC is significantly higher in AD compared to AR for LP/P missense alleles. **(d)** Individual EDC scores for all monogenic genes along with their LP/P frequency shows promising AD gene candidates. All *P* values are indicated in the corresponding figure. All data have been presented as Tukey boxplots, where ‘+’ indicates the mean.

As proof of this association, we performed biophysical mapping of the LB/B and LP/P alleles for 4 monogenic kidney disease genes (*INF2*, *NPHS1*, *ACTN4*, *COL4A3*). In **Figure 6A**, we visualized the mutational patterns of inverted formin-2 (*INF2*), where LP/P missense variants cluster in the N-terminal CBG/FH3 regulatory module (residues 30-233) and LB/B are randomly scattered, respectively. INF2 normally antagonizes diaphanous-related formins (mDia) mediated actin polymerization (Sun et al., 2013), but the clustering of pathogenic variants in the regulatory region causes disinhibition of the INF2-mDia complex in podocytes, and thereby a gain-of-function (GoF) mechanism. Our biophysical postulate for *INF2* is accurate as it independently supports recent *in vivo* knockout studies demonstrating GoF effects from pathogenic missense variants in *INF2* (Subramanian et al., 2024). In contrast to *INF2*, we show the mapping of variants in nephrin (NPHS1) randomly scattered across each Ig-like, C2-type domain following an AR MOI without clustering **(Figure 6B)**. In **Figure 6C**, we demonstrate pathogenic alleles in α-actinin-4, encoded by *ACTN4*, cluster around the CH1-CH2 interface (residues 59-262). Our biophysical data is in accordance with the established GoF mechanism reported by Weins et al., (2007) occurring in the CH1-CH2 domains. Lastly, we mapped alleles in collagen alpha-4 (III) (COL4A3), demonstrating random clustering of LP/P alleles throughout the IDRs of the structure **(Figure 6D)**. Ultimately, in proteins affected by AD inheritance, disease-causing alleles are clustered in important functional domains while those of AR appear to follow a ‘generalized LoF’ mechanism.

**Figure 6.**
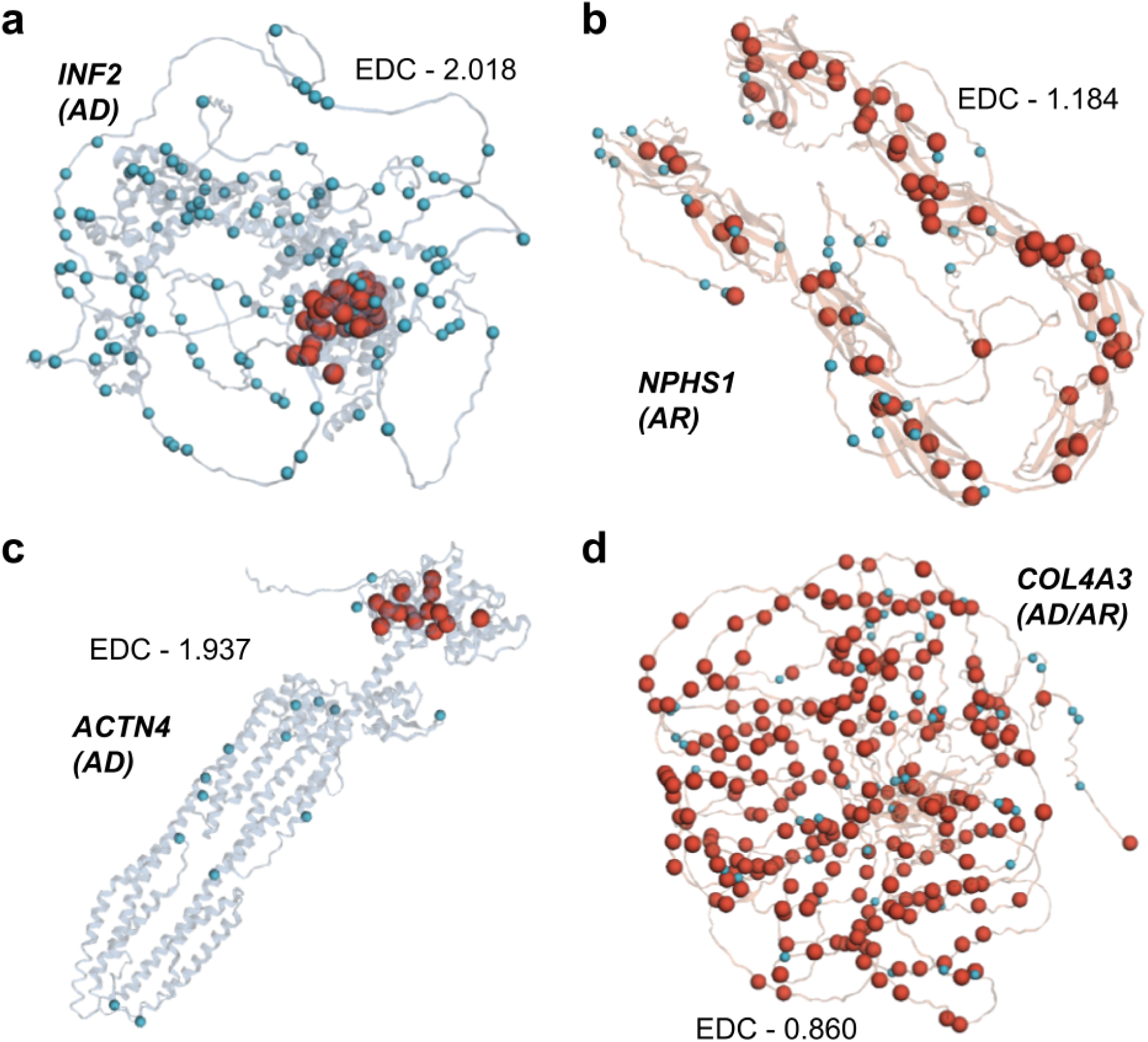
Biophysical mapping studies of missense variants to validate EDC. **(a)** LB/B missense alleles in inverted formin-2 are found throughout the protein structure while LP/P are highly localized in the N-terminal CBG/FH3 regulatory module. (b) In contrast to *INF2*, we show the LB/B and LP/P alleles in nephrin that occur throughout all Ig-like, C2-type domains. **(c)** Similar to *INF2*, we also demonstrate the localization of alleles in AD genes with α-actinin-4, where LP/P alleles are found exclusively in the CH1 and CH2 interface. **(d)** In contrast to all monogenic genes, we show the strong random clustering phenomenon in collagen alpha-4 (III). All LB/B and LP/P missense variants have been shown as light blue and red dots, respectively.

Given the strong predictive power of this genetic data, we sought to predict GoF or dominant negative (DN) in additional AD kidney genes. From the EDC data, we filtered for strong gene candidates by an EDC threshold > 1.6, 10+ genetic variants, and AD MOI, which identified *GATA3*, *LMX1B*, and *PAX2*. In **Figure S6**, we show that these 3 additional independent predictions are accurate and occur through a DN mechanism with extensive experimental evidence.

### Variant effect prediction improves identification of disease-causing variants in autosomal recessive genes and helps broadly downgrade VUSs

Our most ambitious objective of this study was to help address the systemic issue of VUSs, which are dominated by missense variants (*n* = 44,021). Since the 1990s, countless variant effect predictors (VEPs) have been developed and validated to improve the accuracy of binary pathogenicity classification (Riccio et al., 2024; Livesay and Marsh., 2023), with AlphaMissense being the highest performing model to date (auROC ∼ 0.95) (Cheng et al., 2023). Despite their high accuracy, the clinical translation of VEPs remains constrained by *n*-of-1 variant submissions and the absence of disease-specific calibration, despite recent efforts (Badonyi and Marsh., 2026). Critically, while VEPs are exceptional at predicting whether an allele disrupts protein function, structural damage is a necessary but insufficient condition for disease-causing effects. This fundamental aspect of the ‘VUS problem’ remains underappreciated and there is an active need for experimental studies quantifying this discrepancy.

In this study, we first benchmarked AlphaMissense for all classified missense variants across each diagnostic class (*n* = 6,323). The overall auROC was 0.928 (95% CI: 0.920-0.934) for early-onset CKD and the individual classes ranged from 0.901-0.972 **(Table S8)**. Predictions were then obtained for all VUSs, where 23.7% (*n* = 10,442) was pathogenic and 65.1% (*n* = 28,652) was benign. We treated these predictions as the de facto ACMG classifications for VUSs to LP/P and LB/B and quantified their change in variant effects with or without the additional predictions on the clinically validated variant cohorts. If these predictions were accurate in clinical practice, we speculated that the additional variants would strengthen our previous conclusions. First, we calculated the EDC of AR and AD genes to determine spatial clustering effects. In AR genes (*n* = 76), there are no differences in clustering of LP/P (1.08 vs. 1.11; *p* = 0.0608). Unfortunately, in AD genes (*n* = 35), there were significant reductions in clustering (1.28 vs. 1.16; *p* = 0.0461). Though marginal, we performed case studies of our AD candidate genes, *INF2*, *ACTN4*, *GATA3*, *LMX1B*, and *PAX2*, and their individual effects. In **Figure 7A-E**, we demonstrate a major loss of clustering due to the new predictions in all candidates. While pathogenic alleles may be found throughout a protein, not all are necessarily disease-causing. For instance, alleles outside the N-terminal regulatory region of *INF2* will not impose the GoF pathway observed in FSGS. Therefore, our data suggests that VEPs are more suited for AR genes associated with early-onset CKD given their LoF mechanisms. Finally, we defined whether the downgraded VUSs were similar to LB/B variants. In **Figure 7F**, we show that all classes were generally surface-exposed and 25-26% were buried in the structures, non-significant from AM predictions (*p* = 0.2410). Likewise, across each class, 55-59% occurred on loops. These results indicate structural similarity in burial and secondary elements, thereby being important for prioritizing VUSs for molecular resolution. It is our recommendation that VEPs are highly accurate for filtering VUSs that are LB/B missense alleles, but clinical or experimental evidence is required to upgrade these variants to LP/P alleles.

**Figure 7.**
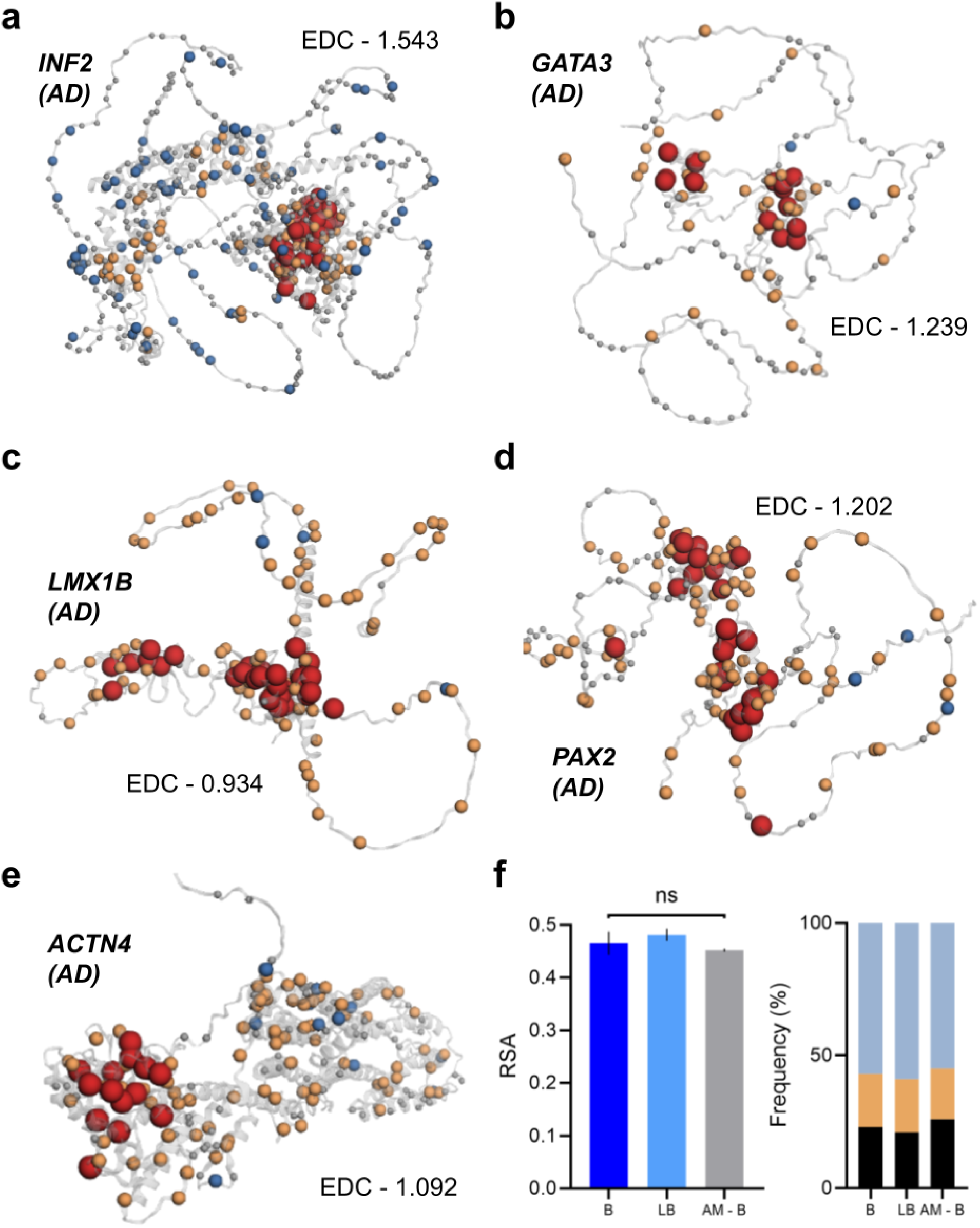
Biophysical mapping studies of AlphaMissense predicted variants. Mapping of LP/P, LB/B, AlphaMissense predicted benign and pathogenic variants onto the structure of **(a)** inverted formin-2, **(b)** GATA binding protein 3, **(c)** LIM Homeobox Transcription Factor 1 Beta, **(d)** Paired box gene 2, **(e)** and α-actinin-4. **(f)** Similarity of biophysical effects in LB, B, and predicted downgraded VUSs in RSA and secondary structural elements. All LB/B, LP/P, VUS downgraded, and VUS upgraded missense variants have been shown as light blue, red, gray, and orange dots. All data have been presented as the mean and 95% CI.

### Proposed standards and guidelines for evaluating gain-of-function and dominant negative effects in monogenic genes causing inherited kidney disease

Lastly, we sought to develop preliminary guidelines for determining GoF and DN effects among all AD genes (*n* = 37) from the NephVar Registry. Previously, we evaluated 6 genes and filtered 19 that lacked sufficient alleles (*n* < 10), leaving 12 unresolved. In the **Figure S7-9** and **Table S9**, we calculated the EDC metric for all 12 remaining AD genes and their molecular effects from reported experimental evidence, respectively. In **Table 1**, we provide suggested standards and guidelines proposed from the NephVar Registry for establishing non-LoF mechanisms in genes causing inherited kidney disease. Our guidelines, inspired by the ACMG criteria, assess the strength of clinical and biophysical evidence supporting a gene. This unified framework has immediate potential for guiding translational research efforts in kidney genetic disorders, in inherited nephrotic syndromes for example. There are two categories for classifying non-LoF variants, including ‘mechanism, very strong’ (MVS) and ‘mechanism strong’ (MS1-3) and two for LoF variants as ‘LoF, strong’ (LS1-3) and ‘LoF, very strong’ (LVS). All criteria are described in detail within **Table 1**.

**Table 1.** Standards and guidelines for non-LoF mechanisms in early-onset CKD proposed from the NephVar Registry. All guidelines have provided examples of monogenic genes evaluated in this study with alignment to ACMG criteria. We suggest two categories for classifying non-LoF variants, including ‘mechanism, very strong’ (MVS) and ‘mechanism strong’ (MS1-3) and two categories for LoF variants as ‘LoF, strong’ (LS1-3) and ‘LoF, very strong’ (LVS).

| **<u>Very strong evidence of non-LoF mechanism</u>** |  | **ACMG** |
| --- | --- | --- |
| MVS | 1. Validated DN or GoF mechanism from functional assays or *in vivo* experimental studies against null LoF variants.<br>Example: *GATA3*, *LMX1B*, *TRPC6*, *INF2*. | PVS1 |
| **<u>Strong evidence of non-LoF mechanism</u>** |  |  |
| MS1 | 1. Distinct genotype-phenotype correlations.<br>Example: *TRPC6*. | PS1-4 |
| MS2 | 1. EDC > 2.000.<br>2. N > 30 LP/P alleles.<br>Example: *INF2*. | PM1-6 |
| MS3 | 1. EDC > 1.200.<br>2. AD MOI validated from segregation analysis.<br>Example: *HNF4A*, *VDR*. | PP1-5,<br>VUS |
| **<u>Strong evidence of LoF mechanism</u>** |  |  |
| LS1 | 1. $1.000 < \text{EDC} < 1.200$<br>2. AR, AR/AD, X-linked MOI from segregation analysis.<br>Example: *COL4A3*, *COL4A4* | PP1-5,<br>VUS |
| LS2 | 1. EDC < 1.000 ('anti-clustering')<br>Example: *COL4A5*. | PM1-6 |
| LS3 | 1. Phenotypic similarity of null LoF to missense variants<br>2. Lack of depletion of LB/B alleles in functional domains<br>Example: *CFI* | PS1-4 |
| **<u>Very strong evidence of LoF mechanism</u>** |  |  |
| LVS | 1. Validated LoF mechanism confirmed for both null LoF and missense variants *in vitro* or *in vivo*.<br>Example: *HNF1B* | PVS1 |

AD, autosomal dominant; AR, autosomal recessive; DN, dominant negative; EDC, extent of disease clustering; MOI, modes of inheritance; PVS, pathogenic very strong; PS, pathogenic strong; PM, pathogenic moderate; PP, pathogenic supporting; VUS, variant of uncertain significance.

The strongest gene candidate supporting our criteria with established personalized care and novel precision medicine approaches is TRPC6 with a GoF mechanism in disease causing variants. In **Figure S7A**, we show disease-causing alleles cluster on the periphery of the cation channel gate despite a lower EDC score, thereby providing a framework for novel drug design and resolving established genotype-phenotype correlations. In a recent observational study, crucial high frequency TRPC6 alleles were identified together with their unique GoF effects causing nephrotic syndrome (McAnallen et al., 2026). Also, a new randomized, placebo-controlled, phase 2 trial of the oral selective TRPC6 inhibitor, BI 764198, showed treatment benefits to patients with GoF TRPC6 alleles (Trachtman et al., 2026). Therefore, our proposed criteria establishing a non-LoF mechanism may be used as a framework to encourage translational research efforts in building targeted therapies with anti-proteinuric effect, with TRPC6 as the driver to translation. Of the remaining 129 kidney disease genes we have studied, INF2 and ACTN4 appear to be the most promising gene candidates due to their known GoF mechanisms and functional localization.

## Discussion

In this study, we introduced the NephVar renal registry, a database that provides clinical and biophysical data for all reported genetic variants in 129 monogenic genes causing early-onset CKD. Using NephVar, we estimated the classification rates of variants in all diagnostic classes and their time to reclassification. Likewise, NephVar revealed the unique disease-causing mechanisms of missense variants, the most frequent variant type in early-onset CKD. Most pathogenic missense variants act through a buried LoF mechanism or alter surface-exposed functional domains. Conversely, for missense alleles in unstructured IDRs, we identified a strong protective association against disease-causing effects. We also demonstrate that the spatial clustering of missense variants is influenced by the MOI, as shown with 5 candidate genes, *INF2*, *ACTN4*, *GATA3*, *LMX1B*, and *PAX2*, with GoF or DN mechanisms. To address the problem of VUSs, we demonstrate that VEPs are accurate in prioritizing LB/B alleles, but fail for LP/P because of their disease-causing effects and MOI. Lastly, we develop standards and guidelines to assess non-LoF mechanisms in genes causing inherited kidney disease. Ultimately, the NephVar platform and its refined resolution of the genetic architecture of early-onset CKD has major implications for clinical diagnostics and translational research in precision nephrology.

Our study underscores the last two decades of progress in kidney genomic medicine. There has been exceptional progress in the identification of genetic variants from NGS methods, but there is not enough data to upgrade or downgrade VUSs, or solve their mechanistic effects. Yet, this issue may not be as severe as expected as 65% of VUSs impose structural effects similar to LB/B variants. This leaves a much smaller fraction of VUSs that could be targeted for scalable mechanistic assays. High quality clinical studies are necessary to refine the disease-causing effects of variants and their respective phenotypes, as shown with the AD genes in this study. For US public policies, our data indicates that the bold predictions by the National Human Genome Research Institute for solving all VUSs by 2030 appears to be improbable in the context of early-onset CKD (Gunter and Green., 2023).

Furthermore, this work emphasizes the priority of experimental studies for AD genes causing early-onset CKD. Though we have strong variant data for 5 candidate genes, most do not have sufficient data to prove a non-LoF mechanism. Recent studies have estimated that 48% of phenotypes in AD genes are caused by GoF or DN effects (Badonyi and Marsh., 2025). It also remains unclear how pathogenic variants in unclustered regions affect phenotypes in AD genes. For instance, what are the mechanistic effects of pathogenic variants downstream of the N-terminal regulatory domain of *INF2*? Of course, the GoF mechanism causing FSGS would likely be absent, but what would be the phenotype in such cases? Deep mutational scanning assays are promising in this regard, but evaluating the diversity of clinical phenotypes would remain a challenge in the field.

Overall, NephVar is the first unified framework to advance precision nephrology efforts in inherited kidney diseases, with a focus on dealing with VUS missense alleles. The establishment of NephVar with early-onset CKD associated disease genes has provided important directions for diagnostic methods and clinical investigations.

## Limitations

There are several limitations of this study. The genetic variant data from ClinVar is dynamic and the quality of submissions are highly variable. Due to the cross-sectional design of this study, the sample sizes and classification rates for all alleles should be interpreted as estimations. While quality control measures were applied to the variant data, even modest changes can alter the sample sizes in all studies reported. To ensure external reproducibility, we archived all ClinVar submissions and released all in-house python scripts. Therefore, we believe that regardless of the variation introduced during sampling, the clinical and biophysical associations identified from this study remain supported for early-onset CKD. Additionally, the genetic variant data in NephVar are gene-level aggregates and do not represent the landscape of any particular phenotype or population. Conversely, in regards to the biophysical studies, we accept the limitation of AlphaFold 2 as an *in silico* prediction. However, many of the proteins analyzed in this study have experimentally resolved structures for important functional domains provided in the Protein Data Bank (PDB). As AlphaFold has been trained on the PDB, predictions for these regions are expected to be highly accurate to experimental resolution. Both of the biophysical and spatial clustering studies depended on the residue geometries in globular domain rather than atomic coordinates, which makes our conclusions robust. We also believe that the frequency of surface-exposed pathogenic alleles may be overestimated in this study due to prediction on monomeric subunits. For instance, it has recently been suggested that podocin, encoded by *NPHS2*, forms an oligomeric state, where surface alleles are buried in the multimeric fold (Mertens et al., 2026).

## Materials and Methods

In this study, we developed the NephVar renal registry, a publicly available database aimed at providing clinical and biophysical data for genetic variants for inherited kidney diseases. Herein, NephVar refined the classification rates and molecular architecture of early-onset CKD from established monogenic genes. This work followed the STROBE guidelines and was exempt from the UCSD Institutional Review Board because the variant data is public and deidentified.

### Establishing the NephVar Renal Registry

The NephVar renal registry is hosted on GitHub Pages, where relevant data for each gene is available in individual files (.html and .csv). All source data and code is publicly available under an MIT license in the GitHub (github.com/NephVar/NephVar). Likewise, the registry is available at the website: nephvar.github.io/NephVar. For the NephVar AI Assistant, we developed a GPT model using training data from the source data provided in the NephVar database. Nephrologists and geneticists may access the NephVar AI Assistant through the landing page of the registry or through OpenAI’s GPT Store as of June 25, 2026: chatgpt.com. We plan to expand the NephVar registry in the near future and will be updating the website and models biannually.

### ClinVar and Disease Classification

NephVar reports variant data for 129 protein-coding genes causing early-onset CKD. All genes and their mode of inheritance have been detailed in Vivante and Hildebrandt., (2016). We collected variant data classified as benign (B), likely benign (LB), variant of unknown significance (VUS), likely pathogenic (LP), and pathogenic (P) according to the American College of Medical Genetics and Genomics and the Association for Molecular Pathology (ACMG/AMP) guidelines (Richards et al., 2015). We excluded variants of conflicting significance in analyses for this study.

### Extraction of ClinVar Variant Datasets

To streamline the process of variant extraction, we initially used the NCBI Entraz API to access ClinVar and download all genetic variants for the 129 genes associated with early-onset CKD. However, we observed discrepancies between the frequency of classified alleles from the API and those manually extracted. Therefore, we manually performed extraction of all genetic variants from ClinVar, obtaining text files for each individual gene stratified by diagnostic class. After extraction, we tabulated all source data and checked for duplications through the ‘VariationID’ variable. Duplicate alleles were separated into their own respective category. All classification methods, reporting criteria, and variables have been described in ClinVar (Langdrum et al., 2018). For classification rates, we used the VariationID from our tabular datasets and matched it with the directory containing each submission record (submission_summary.txt.gz).

### NephVar Variant Cohort

All genetic variants for the 129 monogenic genes were collected from June 1-11, 2026. We have provided the descriptive statistics of the ACMG classification distributions, variant types, and molecular consequences for the cohort in **Table S10-12**.

### AlphaFold and Protein Structure Data

Using the UniProt accession codes for all 129 monogenic genes, we obtained the full-length protein structures of 124 genes using the AlphaFold 2 Structure Database (https://alphafold.ebi.ac.uk/) through the API endpoint (Varadi et al., 2022). Due to the large molecular size of 5 genes (*FREM2*, *FRAS1*, *TNXB*, *FAT1*, and *CUBN*), we used AlphaFold 3 (Abramson et al., 2024) for monomer prediction. Of these AlphaFold 3 predictions, we utilized the highest predicted structure in the NephVar registry. After obtaining all protein structures (.pdb), we used the define secondary structure of proteins (DSSP) algorithm to obtain residue data (Hekkelman et al., 2025). NephVar collected data for the secondary structural elements, accessible surface area, functional annotations, and pLDDT scores for each residue. Using the surface area, we calculated the relative solvent accessibility (RSA) per residue using Tien et al., (2013).

### Molecular Grammars with NARDINI+

Methods for the NARDINI+ algorithm are available at Ruff et al., (2026). Using the database, we obtained the grammars of each gene from their UniProt accession.

### Estimation of Reclassification Rates

The variant dataset stratified by diagnostic class (*n* = 118,367) was used to determine reclassification rates of genetic variants. We set two criteria for including an allele in the estimations. First, a genetic variant is considered reclassified if its latest classification differed from its initial submission from ClinVar regardless of ACMG class. Second, variants with at least a single submission and date of submission were included. Using this dataset, we determined the reclassification rates and the time to reclassification. The time to reclassification was calculated as the difference between the date of the latest and initial submission. Inferential statistics were not performed due to the dataset per diagnostic class not being mutually exclusive.

### Enrichment Analyses of LB/B and LP/P in IDRs

From the 129 monogenic genes, 74 had IDRs in their protein structures reported from the NARDINI+ algorithm. From all genetic variants (*n* = 117,373), we identified 75,643 alleles from these 74 genes. This subcohort of variants was then filtered using the ‘molecular consequence’ label from ClinVar to 32,081 missense variants. The residue position of each variant was then parsed against the IDR Intervals and filtered into non-IDR and IDR subcohorts. We performed enrichment analyses of the LB/B vs. VUS/LP/P and LP/P vs. B/LB/VUS in both cohorts, and further stratified by each diagnostic class. Both statistical models calculated the odds ratio and variance using the Woolf logit method (Agresti., 1999) and Fisher’s exact test (Poisson et al., 2011).

### Solvent Accessibility of Missense Variants

From all missense variants available in NephVar (*n* = 50,425), we obtained the RSA and secondary structure element for each wild-type residue. The variant data was then aggregated and stratified by diagnostic class, then statistical analyses were applied.

### Extent of Disease Clustering

This study modified the EDC metric reported from Gerasimavicius et al., (2022), shown in Eq. (1). For a residue in a monomeric structure, the Cα:Cα distance *D* was computed to all remaining residues with a known allele of interest (LB/B or LP/P) and the closest distance was determined (*D*_min_). The average log distance was computed for the alleles of interest and the remaining residues. EDC was then calculated by dividing the score from the alleles of interest from the remaining residues.

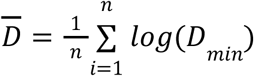

All modes of inheritance for the 129 monogenic genes were obtained from Vivante and Hildebrandt., (2016). EDC was calculated for 85 genes with LP/P alleles and 125 genes for LB/B due to the sample sizes of variants, respectively. When paired for statistical analyses, 82 genes were available in both LP/P and LB/B. To identify strong candidate genes for DN or GoF mechanisms, we searched for genes of AD inheritance, 10+ LP/P alleles, and a score above 1.6.

### Variants of Unknown Significance

We obtained AlphaMissense from Tordai et al., (2024) and Cheng et al., (2023). Predictions were obtained for all VUSs identified from this study (*n* = 44,021), excluding start-stop alleles that are not predicted by the model. After obtaining predictions for all VUSs and treating each prediction as the de facto classification, the EDC metric was applied to assess clustering effects and assess model performance.

## Statistical Analysis

All statistical analyses were performed using an in-house python script with the scipy library or GraphPad Prism. Small sample sizes used the Mann Whitney U-Test and larger samples (*n* > 30) used a Welch’s t-test. DeLong’s test was used to calculate the auROC variance. All data is presented as the mean and 95% confidence intervals. All data files and code is publicly available.

## Data Availability

All data has been provided in the main text and supplemental material. The data has been made freely available at the website(s):

https://nephvar.github.io/NephVar

https://github.com/Joshua-Pillai/NephVar

https://github.com/NephVar/NephVar.

## Acknowledgements

The data reported here have been supplied by ClinVar, a database of the National Center for Biotechnology Information from the National Institutes of Health. The interpretation and reporting of these data are the responsibility of the author(s) and in no way should be seen as an official policy or interpretation of the US government.

## Declarations of interest

J.P.P. has filed a provisional patent application relating to methods for predicting missense variant effects (US Provisional Patent no. 63/768,305).

## Declaration of generative AI

J.P.P. reports the usage of Opus 4.8 by Claude in writing, debugging, and validating the source code. All statistical analyses were manually performed in GraphPad Prism.

## Funding sources

None to declare.

## Author contributions

Conceptualization: J.P.P., J.A.S.; Formal analysis: J.P.P.; Funding acquisition: J.A.S.; Investigation: J.P.P., J.A.S.; Methodology: J.P.P., J.A.S.; Project administration: J.A.S.; Resources: J.P.P., J.A.S.; Supervision: J.A.S.; Validation: J.A.S.; Visualization: J.P.P.; Writing-original draft: J.P.P., J.A.S.; Writing-review & editing: J.P.P., J.A.S.

**#References 39-69 appear in the Supplemental Material.**

## Notes

### Author Declarations

Genetic variant data is available at ClinVar (https://www.ncbi.nlm.nih.gov/clinvar).

