## Supplementary Material for "Resolving the classification rates and molecular architecture of early-onset chronic kidney disease with NephVar"

Pre-print server: medRxiv, CC-BY-NC-ND 4.0.

**Supplemental Information**

**Accurate prediction of gain-of-function or dominant negative mechanisms in 3 AD candidate genes causing early-onset CKD**

Using the NephVar data, we performed biophysical mapping of all LP/P and LB/B alleles in GATA binding protein 3 (GATA3), LIM Homeobox Transcription Factor 1 Beta (LMX1B), and Paired box gene 2 (PAX2), which we have all been predicted to occur through a GoF or DN mechanism from our genetic data. In **Figure S6**, we show the visualizations of all proteins.

In **Figure S6A**, we show the EDC for the GATA binding protein 3 (GATA3), where there are two distinct functional domains clustered, ZnF1 and ZnF2. ZnF1 has been established as a crucial DNA-binding site, where mutations do not cause complete loss of binding, but instead destabilizes localized interactions with FOG2 (Nesbit et al., 2004). Conversely, the disease-causing mechanism for ZnF2 is much more complex. Initial EMSA studies argue against a dominant negative (DN) mechanism of disease (Nesbit et al., 2004), but have reported functional evidence to suggest a DN mechanism for hypoparathyroidism and breast cancer (Ohta et al., 2011; Bianco et al., 2022; El-Assaad et al., 2021). Therefore, our prediction for the non-LoF mechanism using the EDC metric was accurate for *GATA3*.

In **Figure S6B**, we visualize the mapping of LIM Homeobox Transcription Factor 1 Beta (LMX1B), where its variants have pleiotropic effects causing nail-patella syndrome (NPS), nephropathy, and glaucoma. There are two clusters in the homeodomain binding interface, but their mechanisms have remained highly contested. It was suggested that *LMX1B* alleles cause NPS through haploinsufficiency (Dreyer et al., 2000), but there are methodological flaws that do not invalidate a DN effect. Later, Sato et al., (2005) addressed this experimental flaw, confirming the effect and lack of DN. However, this effect was only tested in the promoter region of a single gene, and the broad DN mechanism was not invalidated. In the latest study by Cross et al., (2014) demonstrated a DN effect from a missense variant in mice. Though there is conflicting evidence, *LMX1B* alleles provide some level of evidence for the proposed DN effects.

In **Figure S6C**, we visualize Paired box gene 2 with two clusters that cause an adult-onset of focal segmental glomerulosclerosis. It has been well established that *PAX2* alleles occur through both a DN and haploinsufficiency (Barua et al., 2005; Digvijay et al., 2023; Fletcher et al., 2005). Lastly, we show Wilms tumor 1 (WT1) as a control to demonstrate that not all LP/P alleles cluster in DNA-binding regions of transcription factors. Its molecular mechanism in disease is heterogenous, as both LoF and GoF mechanisms have been observed (Hastie., 2017). Overall, we demonstrate all 3 gene candidates have disease mechanisms that may occur through a non-LoF effect and validate our independent prediction of missense alleles in these genes. In **Table S7**, we provided the postulated mechanisms for all 37 AD genes.

**Gene screening panels of early-onset CKD in NephVar**

This gene list is adapted from Vivante and Hildebrandt., (2016).

CAKUT - *BMP4* (AD), *CHD1L* (AD), *DSTYK* (AD), *EYA1* (AD), *GATA3* (AD), *HNF1B* (AD), *MUC1* (AD), *PAX2* (AD), *RET* (AD), *ROBO2* (AD), *SALL1* (AD), *SIX1* (AD), *SIX2* (AD), *SIX5* (AD), *SOX17* (AD), *SRGAP1* (AD), *TBX18* (AD), *TNXB* (AD), *UMOD* (AD), *UPK3A* (AD), *WNT4* (AD), *ACE* (AR), *AGT* (AR), *AGTR1* (AR), *CHRM3* (AR), *FGF20* (AR), *FRAS1* (AR), *FREM1* (AR), *FREM2* (AR), *GRIP1* (AR), *HPSE2* (AR), *ITGA8* (AR), *LRIG2* (AR), *REN* (AR), *TRAP1* (AR), and *KAL1* (X-linked).

SRNS - *ADCK4* (AR), *ARHGDIA* (AR), *CD2AP* (AR), *CFH* (AR), *COQ2* (AR), *COQ6* (AR), *CRB2* (AR), *CUBN* (AR), *DGKE* (AR), *EMP2* (AR), *FAT1* (AR), *ITGA3* (AR), *ITGB4* (AR), *KANK1* (AR), *KANK2* (AR), *KANK4* (AR), *LAMB2* (AR), *MTTL1* (AR), *MYO1E* (AR), *NPHS1* (AR), *NPHS2* (AR), *NUP93* (AR), *NUP107* (AR), *NUP205* (AR), *PDSS2* (AR), *PLCE1* (AR), *PTPRO* (AR), *SCARB2* (AR), *SMARCAL1* (AR), *WDR73* (AR), *XPO5* (AR), *ACTN4* (AD), *ANLN* (AD), *ARHGAP24* (AD), *INF2* (AD), *LMX1B* (AD), *MYH9* (AD), *TRPC6* (AD), and *WT1* (AD).

CGN - *CFH* (AR), *CFI* (AD), *CFHR5* (AD), *FN1* (AD), *COL4A3* (AD/AR), *COL4A4* (AD/AR), *CD46* (AD/AR), *C3* (AD/AR), *COL4A5* (X-linked), and *COL4A6* (X-linked).

NPHP - *NPHP1* (AR), *INVS* (AR), *NPHP3* (AR), *NPHP4* (AR), *IQCB1* (AR), *CEP290* (AR), *GLIS2* (AR), *RPGRIP1L* (AR), *NEK8* (AR), *SDCCAG8* (AR), *TMEM67* (AR), *TTC21B* (AR), *WDR19* (AR), *ZNF423* (AR), *CEP164* (AR), and *ANKS6* (AR).

USD - *ADCY10* (AD), *AGXT* (AR), *APRT* (AR), *ATP6V0A4* (AR), *ATP6V1B1* (AR), *CA2* (AR), *CASR* (AD), *CLCN5* (XR), *CLCNKB* (AR), *CLDN16* (AR), *CLDN19* (AR), *CYP24A1* (AR), *FAM20A* (AR), *GRHPR* (AR), *HNF4A* (AD), *HOGA1* (AR), *HPRT1* (XR), *KCNJ1* (AR), *OCRL* (XR), *SLC12A1* (AR), *SLC22A12* (AD/AR), *SLC2A9* (AD/AR), *SLC34A1* (AD/AR), *SLC34A3* (AR), *SLC3A1* (AR), *SLC4A1* (AD/AR), *SLC7A9* (AD/AR), *SLC9A3R1* (AD), *VDR* (AD), and *XDH* (AR).

Autosomal dominant (AD), autosomal recessive (AR), autosomal dominant or recessive (AD/AR), and X-linked.

**Supplemental Figure Description**

**Table S1.** Trends in reclassification rates of variants with at least 2 submissions. P: pathogenic; LP: likely pathogenic; VUS: variant of unknown significance; LB: likely benign; B: benign. 'Latest' indicates the most recent reclassification of a genetic variant.

**Table S2.** Secondary structural elements for each diagnostic class stratified by ACMG classifications for loops, sheets, and helix. USD: Urinary stone disease; SRNS: steroid-resistant nephrotic syndrome; CAKUT: congenital anomalies of the kidney and urinary tract; CGN: chronic glomerulonephritis; NPHP: nephronophthisis-related ciliopathies.

**Table S3.** Burial states (RSA) of all ACMG classes stratified by diagnostic classes. Results from Mann-Whitney U-Test comparing P/LP vs. B/LB for each group.

**Table S4.** Position of surface-exposed pathogenic missense variants. Location may be in IDRs, relevant functional domains, or functional class. IDRs: intrinsically disordered regions.

**Table S5.** Distribution of ACMG classified intronic variants, where 75.43% of alleles are classified as LB/B.

**Table S6.** Distribution of ACMG classified synonymous variants, where 90.80% of all alleles are classified as LB/B.

**Table S7.** Distribution of ACMG classified frameshift variants, where 80.81% of alleles have been classified as LP/P.

**Table S8.** Performance of AlphaMissense on classified genetic variants in early-onset CKD stratified by diagnostic classes with the area under the ROC curve (auROC).

**Table S9.** Mechanistic effects of AD monogenic genes causing early-onset CKD. HI: Haploinsufficiency; GOF: gain-of-function; DN: dominant negative.

**Table S10.** Distribution of ACMG classification from the NephVar cohort. Genetic variants defined as 'Other' are labels outside of the ACMG/AMP guidelines, such as risk factors, borderline categories (LP/P; VUS/LP, etc).

**Table S11.** Distribution of variant types from the NephVar cohort, where most are single nucleotide variants.

**Table S12.** Distribution of molecular consequences of genetic variants from the NephVar cohort. This table represents the 15 highest reported consequences.

**Figure S1. Homepage of Clinical Data for Genetic Variants. (a)** Example of *COL4A3* genetic variants, reclassified variants. **(b)** After tapping on an individual variant, users may observe the submission history, phenotypes, and the Variation ID for ClinVar. The example of *COL4A5* G624D is provided as the most submitted variant in CGN and the second greatest for *COL4A3* Q1495R.

**Figure S2. Homepage of Biophysical Analyses. (a)** Biophysical analyses may be performed for the molecular grammars or structural data of a given protein. **(b)** Example of structural data for the *CFHR5* gene, which includes functional annotations, structural elements, relative solvent accessibility (RSA), domains, and confidence (pLDDT).

**Figure S3. Conversation with the NephVar AI Assistant.** From Al Riyami et al., (2026), we prompted the model to predict the pathogenicity of 3 VUSs that have been identified in Omani patients with nephrotic syndrome.

**Figure S4. Challenges of the NephVar AI Assistant in accessing variant data from the NephVar registry.** The AI model fails to identify the presence of two SRNS gene variants in NephVar although present in the dataset.

**Figure S5. Confidence of AlphaFold 2 monomeric predictions.** Of all 129 structures, 95 were above the mean threshold of 70%, indicating high confidence prediction.

**Figure S6. EDC of autosomal dominant candidate genes.** EDC of the **(a)** GATA binding protein 3, **(b**) LIM Homeobox Transcription Factor 1 Beta, **(c)** and Paired box gene 2, all indicating high clustering of LP/P alleles. **(d)** Wilms Tumor 1 was used as a control to show transcription factors do not all cluster in DNA-binding regions and that the proteins shown in panel (a-b) are unique.

**Figure S7. EDC of autosomal dominant genes.** EDC of the **(a)** Transient Receptor Potential Canonical 6, **(b**) calcium-sensing receptor, and **(c)** Hepatocyte Nuclear Factor 4 Alpha, all indicating high clustering of LP/P alleles.

**Figure S8. EDC of autosomal dominant genes.** EDC of the **(a)** Rearranged during Transfection, **(b**) Myosin Heavy Chain 9, **(c)** Hepatocyte Nuclear Factor 1 Homeobox B, **(d)** and complement factor 1 all indicating high clustering of LP/P alleles.

**Figure S9. EDC of autosomal dominant genes.** EDC of the **(a)** Vitamin D Receptor, **(b**) Sine oculis homeobox homolog 1, **(c)** Eyes Absent Homolog 1, **(d)** Fibronectin 1, and **(e)** Uromodulin all indicating high clustering of LP/P alleles.

Table S1. Pillai and Sayer., (2026).

| First | Latest P | Latest LP | Latest VUS | Latest LB | Latest B |
|---|---|---|---|---|---|
| P | 1,083 | 682 | 35 | 3 | 0 |
| LP | 699 | 903 | 64 | 1 | 1 |
| VUS | 37 | 67 | 12,893 | 191 | 50 |
| LB | 0 | 0 | 177 | 4,767 | 1,319 |
| B | 0 | 0 | 2 | 309 | 2,214 |

Table S2. Pillai and Sayer., (2026).

| Cohort | ACMG | *n* | Helix | Sheet | Loop |
|---|---|---|---|---|---|
| CGN | P | 256 | 0.4% | 7.4% | 92.2% |
| CGN | LP | 1,240 | 1.1% | 7.3% | 91.6% |
| CGN | VUS | 5,189 | 5.9% | 26.2% | 67.8% |
| CGN | LB | 343 | 2.0% | 20.1% | 77.8% |
| CGN | B | 59 | 0.0% | 13.6% | 86.4% |
| CAKUT | P | 148 | 30.4% | 31.8% | 37.8% |
| CAKUT | LP | 294 | 27.9% | 24.5% | 47.6% |
| CAKUT | VUS | 11,984 | 16.0% | 29.1% | 54.9% |
| CAKUT | LB | 629 | 11.9% | 31.8% | 56.3% |
| CAKUT | B | 182 | 15.4% | 31.9% | 52.7% |
| SRNS | P | 144 | 36.8% | 23.6% | 39.6% |
| SRNS | LP | 362 | 37.3% | 20.7% | 42.0% |
| SRNS | VUS | 11,787 | 25.8% | 18.2% | 56.1% |
| SRNS | LB | 824 | 25.2% | 15.0% | 59.7% |
| SRNS | B | 211 | 25.1% | 20.4% | 54.5% |
| USD | P | 304 | 44.4% | 16.4% | 39.1% |
| USD | LP | 596 | 48.3% | 14.1% | 37.6% |
| USD | VUS | 6,955 | 46.8% | 12.0% | 41.2% |
| USD | LB | 278 | 42.8% | 11.5% | 45.7% |
| USD | B | 78 | 37.2% | 10.3% | 52.6% |
| NPHP | P | 43 | 27.9% | 34.9% | 37.2% |
| NPHP | LP | 117 | 34.2% | 25.6% | 40.2% |
| NPHP | VUS | 8,106 | 47.2% | 10.2% | 42.6% |
| NPHP | LB | 156 | 41.0% | 9.6% | 49.4% |

Table S3. Pillai and Sayer., (2026).

| Cohort | Class | *n* | Median | % Buried | *p*-value* |
|---|---|---|---|---|---|
| CGN | P/LP | 19 | 0.712 | 10.5% | 2.8240e-01 |
| CGN | B/LB | 13 | 0.771 | 7.7% | |
| CAKUT | P/LP | 7 | 0.236 | 57.1% | 4.1802e-01 |
| CAKUT | B/LB | 19 | 0.337 | 36.8% | |
| SRNS | P/LP | 21 | 0.107 | 66.7% | 4.5893e-03 |
| SRNS | B/LB | 24 | 0.456 | 33.3% | |
| USD | P/LP | 40 | 0.128 | 62.5% | 4.7274e-03 |
| USD | B/LB | 19 | 0.403 | 31.6% | |
| NPHP | P/LP | 4 | 0.299 | 50.0% | 5.4874e-01 |
| NPHP | B/LB | 13 | 0.416 | 30.8% | |

Table S4. Pillai and Sayer., (2026).

| **Category** | ***n*** | **%** |
|---|---|---|
| domain only | 196 | 25.5% |
| IDR only | 27 | 3.5% |
| Functional class only | 109 | 14.2% |
| Domain + functional class | 18 | 2.3% |
| Domain + IDR | 1 | 0.1% |
| IDR + functional class | 0 | 0.0% |
| Domain + IDR + functional class | 0 | 0.0% |
| None | 418 | 54.4% |

Table S5. Pillai and Sayer., (2026).

| Panel | P | LP | VUS | LB | B | Other | Total | % |
|---|---|---|---|---|---|---|---|---|
| CGN | 34 | 41 | 820 | 2,670 | 875 | 142 | 4,582 | 16.4 |
| CAKUT | 58 | 65 | 1,441 | 2,987 | 971 | 204 | 5,726 | 20.5 |
| SRNS | 86 | 92 | 1,554 | 4,382 | 1,814 | 228 | 8,156 | 29.2 |
| USD | 71 | 98 | 758 | 2,320 | 1,024 | 112 | 4,383 | 15.7 |
| NPHP | 55 | 58 | 837 | 3,396 | 626 | 107 | 5,079 | 18.2 |
| All | 304 | 354 | 5,410 | 15,755 | 5,310 | 793 | 27,926 | 100.0 |

Table S6. Pillai and Sayer., (2026).

| Panel | P | LP | VUS | LB | B | Other | Total | % |
|---|---|---|---|---|---|---|---|---|
| CGN | 3 | 2 | 155 | 2,741 | 129 | 112 | 3,142 | 12.8 |
| CAKUT | 1 | 2 | 277 | 5,391 | 309 | 385 | 6,365 | 25.9 |
| SRNS | 2 | 2 | 294 | 5,611 | 386 | 348 | 6,643 | 27.1 |
| USD | 2 | 7 | 199 | 3,343 | 155 | 164 | 3,870 | 15.8 |
| NPHP | 5 | 1 | 203 | 4,147 | 67 | 92 | 4,515 | 18.4 |
| All | 13 | 14 | 1,128 | 21,233 | 1,046 | 1,101 | 24,535 | 100.0 |

Table S7. Pillai and Sayer., (2026).

| Panel | P | LP | VUS | LB | B | Other | Total | % |
|---|---|---|---|---|---|---|---|---|
| CGN | 447 | 296 | 43 | 0 | 0 | 134 | 920 | 18.9 |
| CAKUT | 529 | 241 | 99 | 3 | 0 | 59 | 931 | 19.1 |
| SRNS | 410 | 293 | 111 | 1 | 2 | 98 | 915 | 18.8 |
| USD | 527 | 296 | 43 | 1 | 0 | 88 | 955 | 19.6 |
| NPHP | 593 | 306 | 48 | 1 | 1 | 203 | 1,152 | 23.6 |
| All | 2,506 | 1,432 | 344 | 6 | 3 | 582 | 4,873 | 100.0 |

Table S8. Pillai and Sayer., (2026).

| **Panel** | **auROC** | **95% CI** | ***n*** | **P/LP** | **B/LB** |
|---|---|---|---|---|---|
| Overall | 0.9276 | 0.9206–0.9340 | 6,323 | 3,503 | 2,820 |
| CGN | 0.9720 | 0.9617–0.9808 | 1,898 | 1,496 | 402 |
| CAKUT | 0.9265 | 0.9117–0.9405 | 1,251 | 441 | 810 |
| SRNS | 0.9135 | 0.8983–0.9274 | 1,541 | 506 | 1,035 |
| USD | 0.9009 | 0.8816–0.9198 | 1,256 | 900 | 356 |
| NPHP | 0.9013 | 0.8683–0.9301 | 377 | 160 | 217 |

Table S9. Pillai and Sayer., (2026).

| Gene | Genetic Mechanism | Evidence | Structure |
|---|---|---|---|
| *ACTN4* | GoF | Aforementioned | AlphaFold 2 |
| *ADCY10* | Unknown | *n* < 10 for LP/P | NA |
| *ANLN* | Unknown | *n* < 10 for LP/P | NA |
| *ARHGAP24* | Unknown | *n* < 10 for LP/P | NA |
| *BMP4* | Unknown | *n* < 10 for LP/P | NA |
| *CASR* | GoF/LoF | Hannan et al., (2013) | Gao et al., (2021) |
| *CFHR5* | Unknown | *n* < 10 for LP/P | NA |
| *CFI* | LoF | Kavanagh et al., (2015) | AlphaFold 3 |
| *CHD1L* | Unknown | *n* < 10 for LP/P | NA |
| *DSTYK* | Unknown | *n* < 10 for LP/P | NA |
| *EYA1* | GoF/LoF | Wong et al., (2012) | AlphaFold 2 |
| *FN1* | GoF | Castelletti et al., (2008) | AlphaFold 3 |
| *GATA3* | DN/HI | Aforementioned | AlphaFold 2 |
| *HNF1B* | HI | Niborski et al., (2021) | Lu et al., (2007) |
| *HNF4A* | GoF | DeForest et al., (2023) | Chandra et al., (2013) |
| *INF2* | GoF | Aforementioned | AlphaFold 2 |
| *LMX1B* | DN/HI | Aforementioned | AlphaFold 2 |
| *MUC1* | Unknown | *n* < 10 for LP/P | NA |
| *MYH9* | DN | De Rocco et al., (2013) | Casas-Mao et al., (2026) |
| *PAX2* | DN/HI | Aforementioned | AlphaFold 2 |
| *RET* | LoF | Shi et al., (2025) | Li et al., (2019) |
| *ROBO2* | Unknown | *n* < 10 for LP/P | NA |
| *SALL1* | Unknown | *n* < 10 for LP/P | NA |

| | | | |
|---|---|---|---|
| *SIX1* | HI | Ruf et al., (2026) | Patrick et al., (2013) |
| *SIX2* | Unknown | *n* < 10 for LP/P | NA |
| *SIX5* | Unknown | *n* < 10 for LP/P | NA |
| *SLC9A3R1* | Unknown | *n* < 10 for LP/P | NA |
| *SOX17* | Unknown | *n* < 10 for LP/P | NA |
| *SRGAP1* | Unknown | *n* < 10 for LP/P | NA |
| *TBX18* | Unknown | *n* < 10 for LP/P | NA |
| *TNXB* | Unknown | *n* < 10 for LP/P | NA |
| *TRPC6* | GoF/LoF | McAnallen et al., (2026) | Tang et al., (2019) |
| *UMOD* | GoF | Econimo et al., (2022) | Stsiapanava et al., (2022) |
| *UPK3A* | Unknown | *n* < 10 for LP/P | NA |
| *VDR* | DN | Feldman et al., (2014) | Sigueiro et al., (2022) |
| *WNT4* | Unknown | *n* < 10 for LP/P | NA |
| *WT1* | GoF/LoF | Aforementioned | AlphaFold 2 |

Table S10. Pillai and Sayer., (2026).

| **ACMG** | ***n*** |
|---|---|
| VUS | 52,130 |
| LB | 37,263 |
| P | 9,400 |
| B | 7,702 |
| LP | 6,376 |
| Other | 4,502 |

Table S11. Pillai and Sayer., (2026).

| **Variant type** | ***n*** |
|---|---|
| Single Nucleotide Variant | 102,223 |
| Deletion | 6,102 |
| Duplication | 2,617 |
| Copy Number Loss | 2,072 |
| Copy Number Gain | 1,955 |
| Microsatellite | 1,186 |
| Indel | 651 |
| Insertion | 420 |
| Inversion | 81 |
| Haplotype | 27 |
| Variation | 21 |
| Complex | 16 |
| Translocation | 1 |
| Phase unknown | 1 |

Table S12. Pillai and Sayer., (2026).

| **Molecular genetic consequence** | ***n*** |
|---|---|
| missense variant | 39,644 |
| intron variant | 21,780 |
| synonymous variant | 19,704 |
| missense variant \| non-coding transcript variant | 4,884 |
| frameshift variant | 4,066 |
| nonsense | 2,793 |
| missense variant \| intron variant | 2,139 |
| 3 prime UTR variant | 2,023 |
| synonymous variant \| non-coding transcript variant | 1,930 |
| splice donor variant | 1,434 |
| synonymous variant \| intron variant | 1,310 |
| splice acceptor variant | 1,052 |
| non-coding transcript variant \| missense variant | 684 |
| missense variant \| non-coding transcript variant \| intron variant | 574 |

*Note: this is an incomplete list, and is the highest 15 categories reported to date.

Figure S1. Pillai and Sayer., (2026).

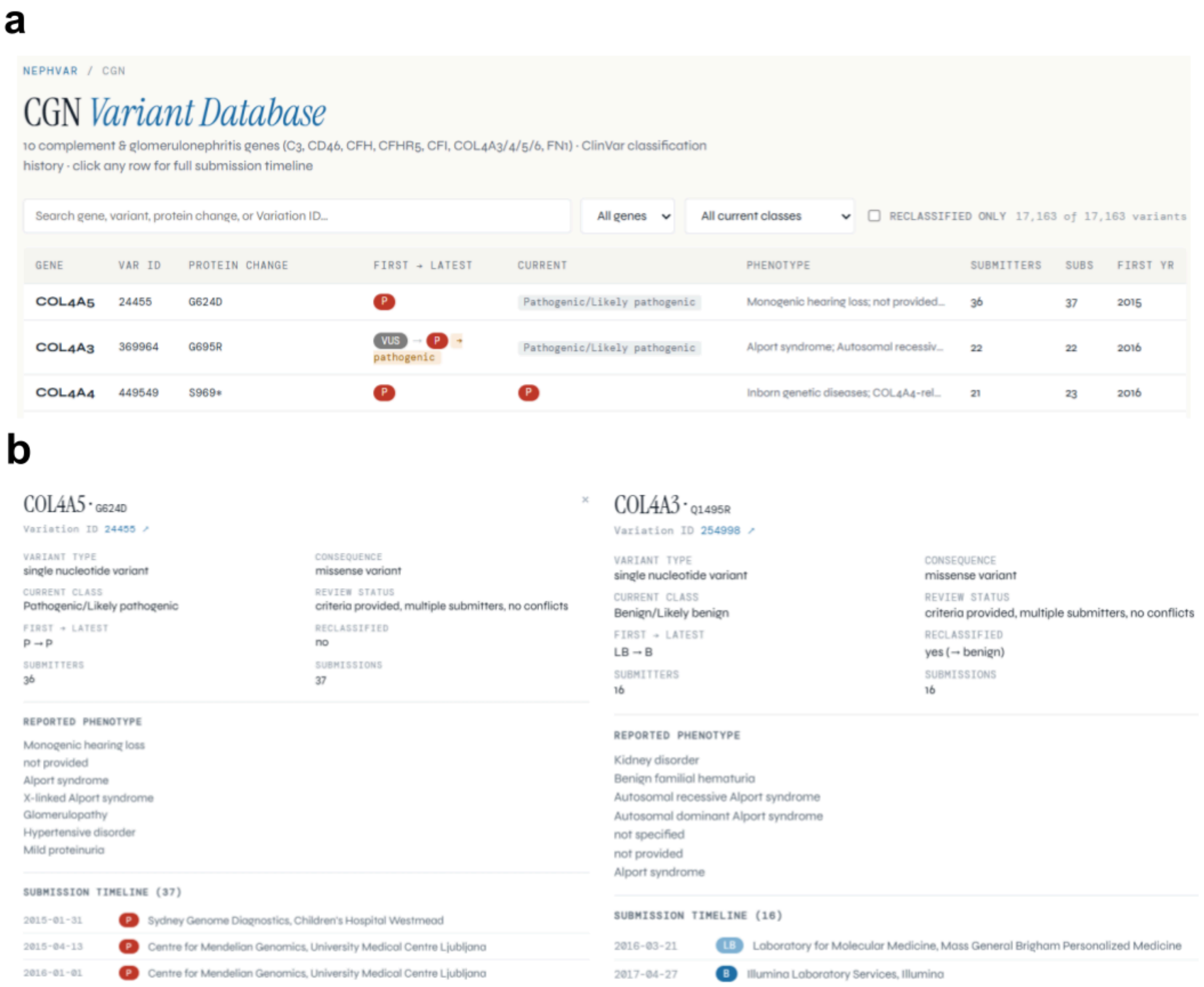

Figure S2. Pillai and Sayer., (2026).

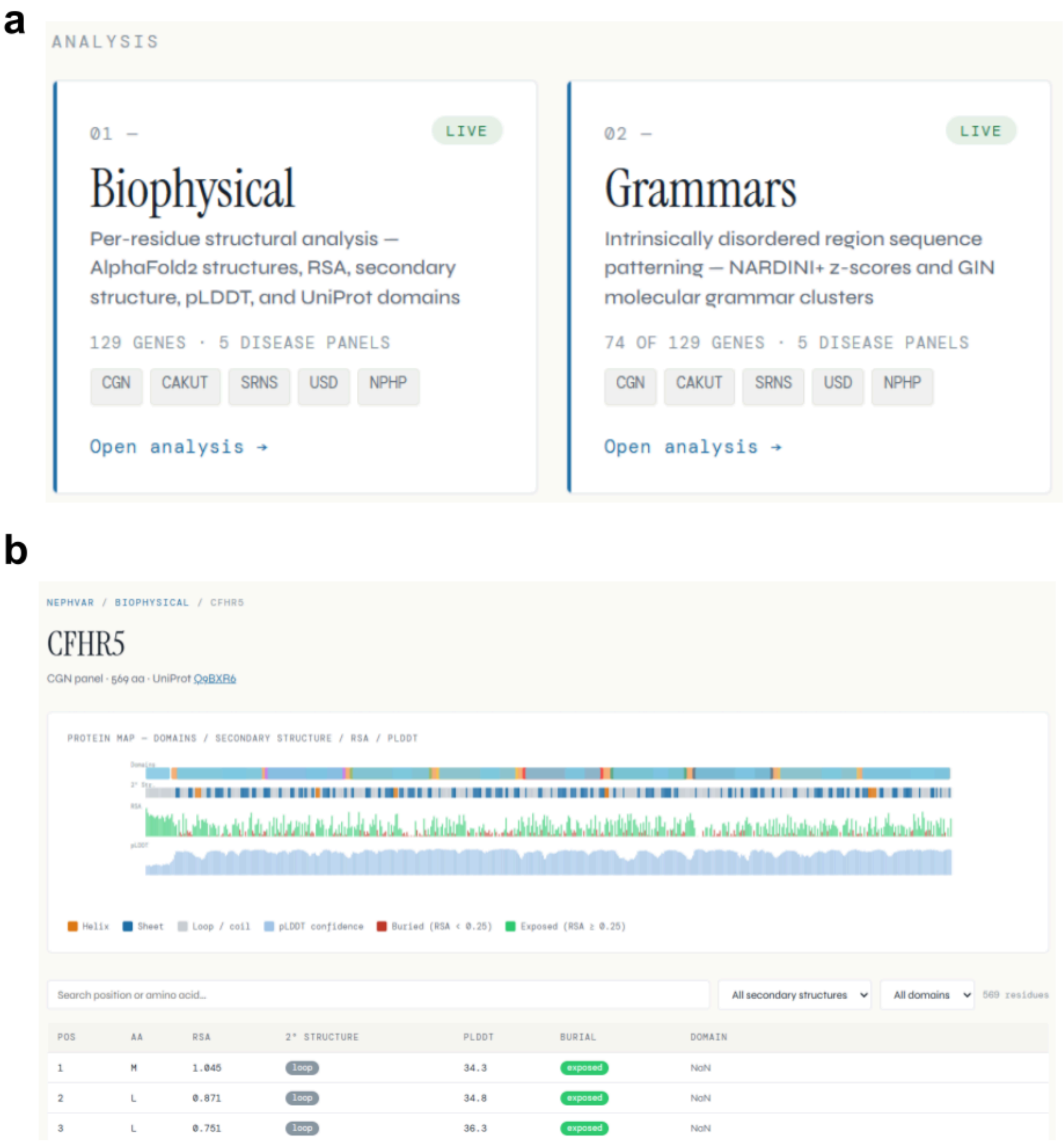

Figure S3. Pillai and Sayer., (2026).

Hi NephVar, hope you are doing great! I am the chief nephrologist at an academic medical center in the United States, and you are my senior research scientist. Using NephVar, could you please give me all clinical and biophysical data available for the following alleles from my cohort with nephrotic syndrome?
DGKE; p.(Lys109Glu)
CD2AP; p.(Arg111His)
SMARCAL1; p.(Lys849Asn)

I'll cross-check the local NephVar variant databases first, then verify the public NephVar/GitHub context for any fields or interpretation that aren't obvious from the CSVs.

Thought for 1m 15s ›

I screened the **NephVar SRNS database** because all three genes are in the steroid-resistant nephrotic syndrome panel; the public NephVar SRNS page lists **CD2AP, DGKE, and SMARCAL1** among the ClinVar-derived SRNS genes and reports **30,692 unique SRNS variants** total. nephvar.github.io NephVar's biophysical module is intended to provide **AlphaFold2 structure, DSSP secondary structure, RSA, pLDDT, and UniProt domain features**, but the three gene pages currently [illegible]play only domain/filter metadata and no residue rows for direct RSA/pLDDT extraction. nephvar.github.io +3

Thought for 58s ›

My suggested **research reclassification**, using NephVar plus currently visible external data, would be:

| Allele | I would call it, beyond VUS | Confidence | Rationale |
|---|---|---|---|
| **DGKE p.(Lys109Glu), K109E** | **Likely pathogenic**, especially if **biallelic/homozygous** and phenotype includes early-onset nephrotic syndrome, TMA/aHUS, MPGN/C3G, or proteinuria | **Moderate** | Reported in DGKE nephropathy literature as **p.K109E/p.K109E**; supplemental prediction data call K109E damaging/deleterious by multiple tools. NephVar currently lists it as VUS, but the literature signal is stronger than the current ClinVar-style assertion. PMC +1 |
| **CD2AP p.(Arg111His), R111H** | **Likely benign / risk-modifier at most**, not monogenic causal by itself | **Low–moderate** | NephVar/ClinVar lists VUS with limited clinical evidence; ClinVarMiner reports rs192679464 with gnomAD frequency around **0.00024**, which feels high for a fully penetrant rare monogenic nephrotic syndrome allele. The protein context is interesting, but there is not enough disease evidence. NCBI +1 |
| **SMARCAL1 p.(Lys849Asn), K849N** | **Likely pathogenic only if biallelic/in trans with another pathogenic SMARCAL1 allele and SIOD-compatible phenotype; otherwise VUS-favor pathogenic** | **Low–moderate** | Exact variant is absent from local NephVar tables but has now been reported as **SMARCAL1 c.2547G>T, p.Lys849Asn** in a child with Schimke immuno-osseous dysplasia, where it was still described as VUS. SMARCAL1 has definitive autosomal-recessive disease validity for SIOD, and the residue is in the C-terminal helicase-region neighborhood. PMC +1 |

Figure S4. Pillai and Sayer., (2026).

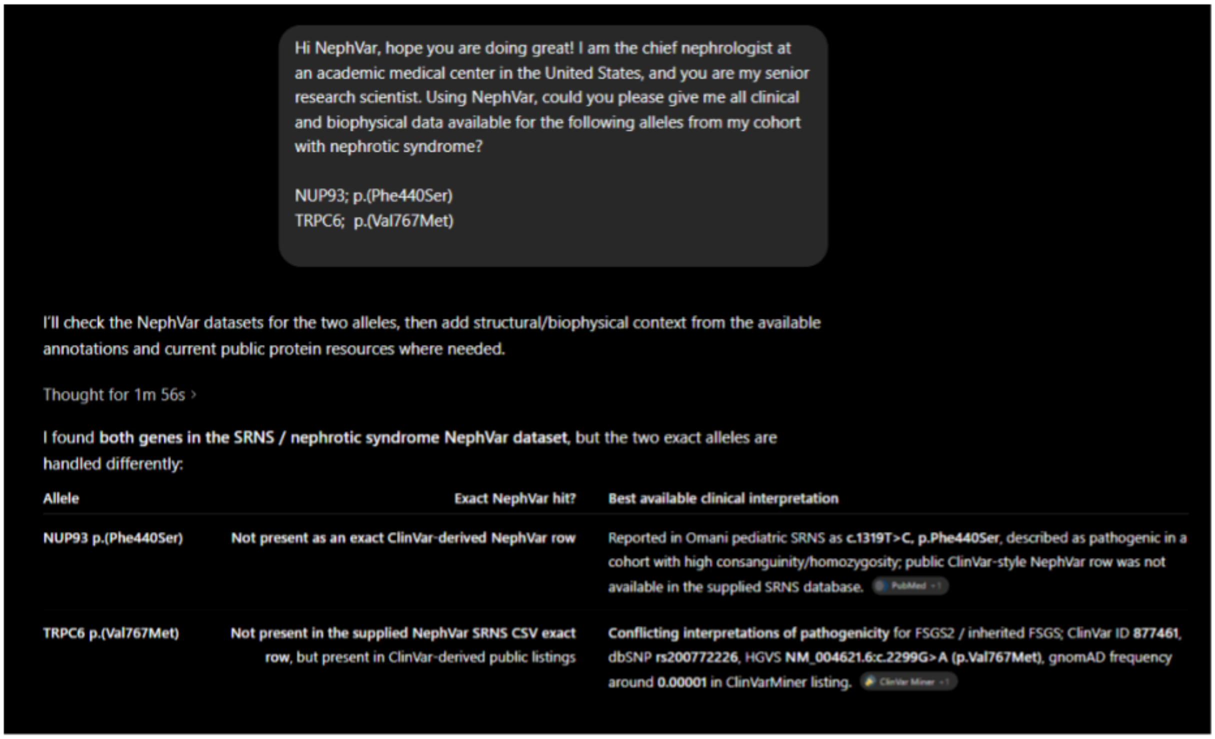

Hi NephVar, hope you are doing great! I am the chief nephrologist at an academic medical center in the United States, and you are my senior research scientist. Using NephVar, could you please give me all clinical and biophysical data available for the following alleles from my cohort with nephrotic syndrome?

NUP93; p.(Phe440Ser)
TRPC6;  p.(Val767Met)

I'll check the NephVar datasets for the two alleles, then add structural/biophysical context from the available annotations and current public protein resources where needed.

Thought for 1m 56s ›

I found **both genes in the SRNS / nephrotic syndrome NephVar dataset**, but the two exact alleles are handled differently:

| Allele | Exact NephVar hit? | Best available clinical interpretation |
|---|---|---|
| **NUP93 p.(Phe440Ser)** | **Not present as an exact ClinVar-derived NephVar row** | Reported in Omani pediatric SRNS as **c.1319T>C, p.Phe440Ser**, described as pathogenic in a cohort with high consanguinity/homozygosity; public ClinVar-style NephVar row was not available in the supplied SRNS database. |
| **TRPC6 p.(Val767Met)** | **Not present in the supplied NephVar SRNS CSV exact row**, but present in ClinVar-derived public listings | **Conflicting interpretations of pathogenicity** for FSGS2 / inherited FSGS; ClinVar ID **877461**, dbSNP **rs200772226**, HGVS **NM_004621.6:c.2299G>A (p.Val767Met)**, gnomAD frequency around **0.00001** in ClinVarMiner listing. |

Figure S5. Pillai and Sayer., (2026).

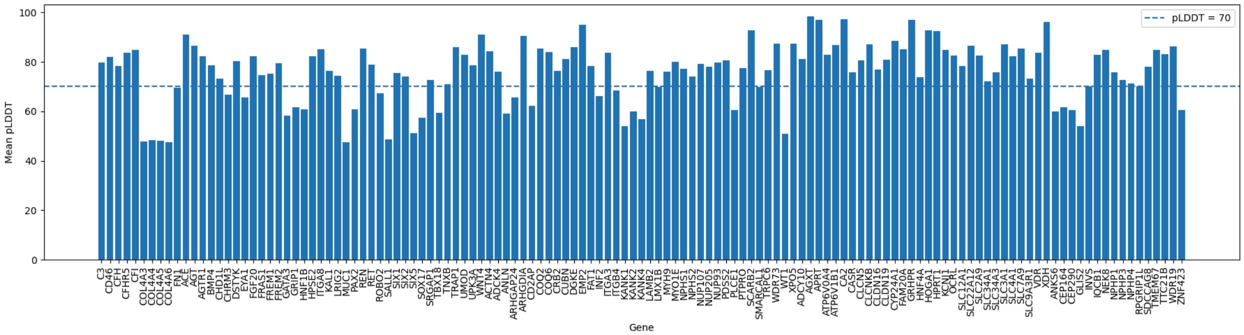

Figure S6. Pillai and Sayer., (2026).

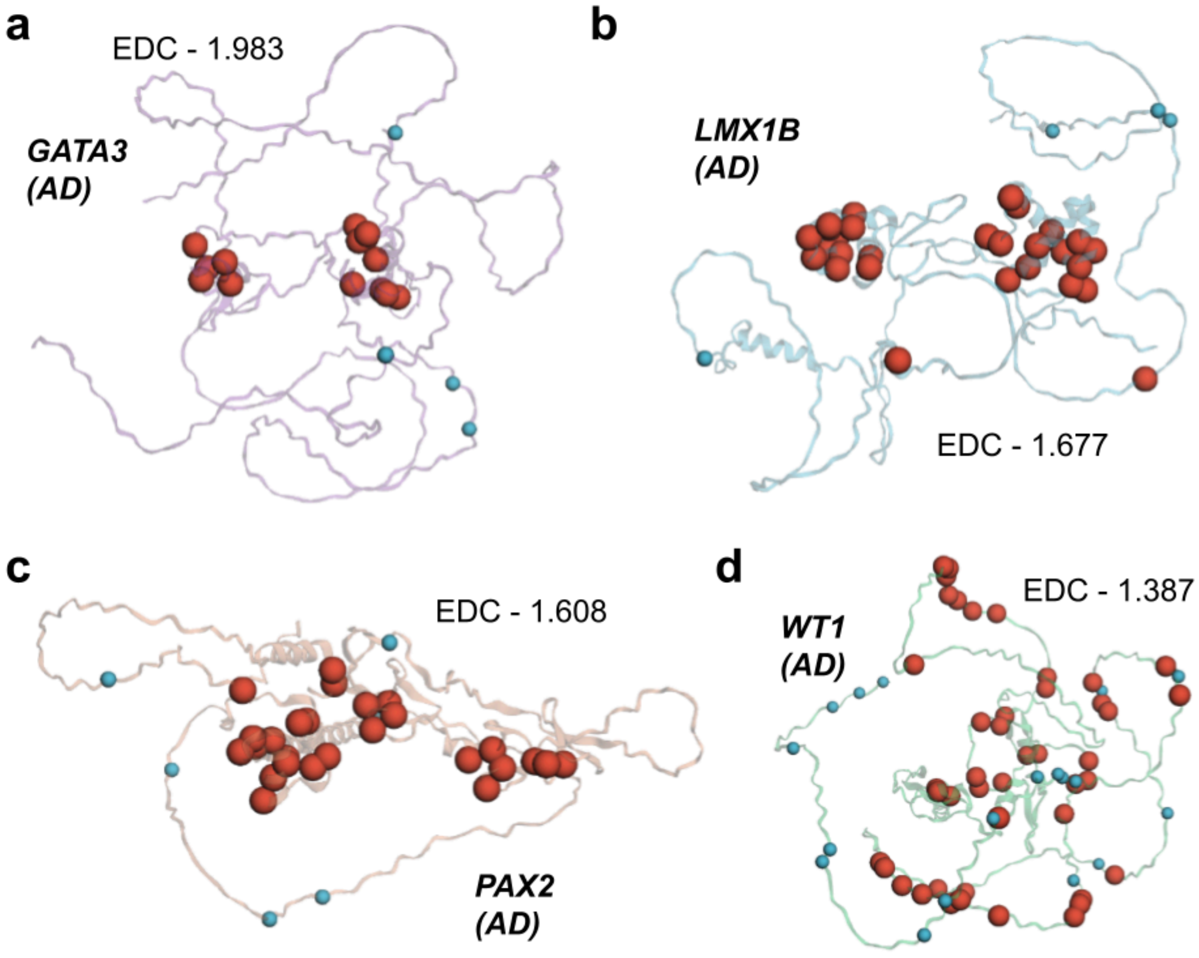

Figure S7. Pillai and Sayer., (2026).

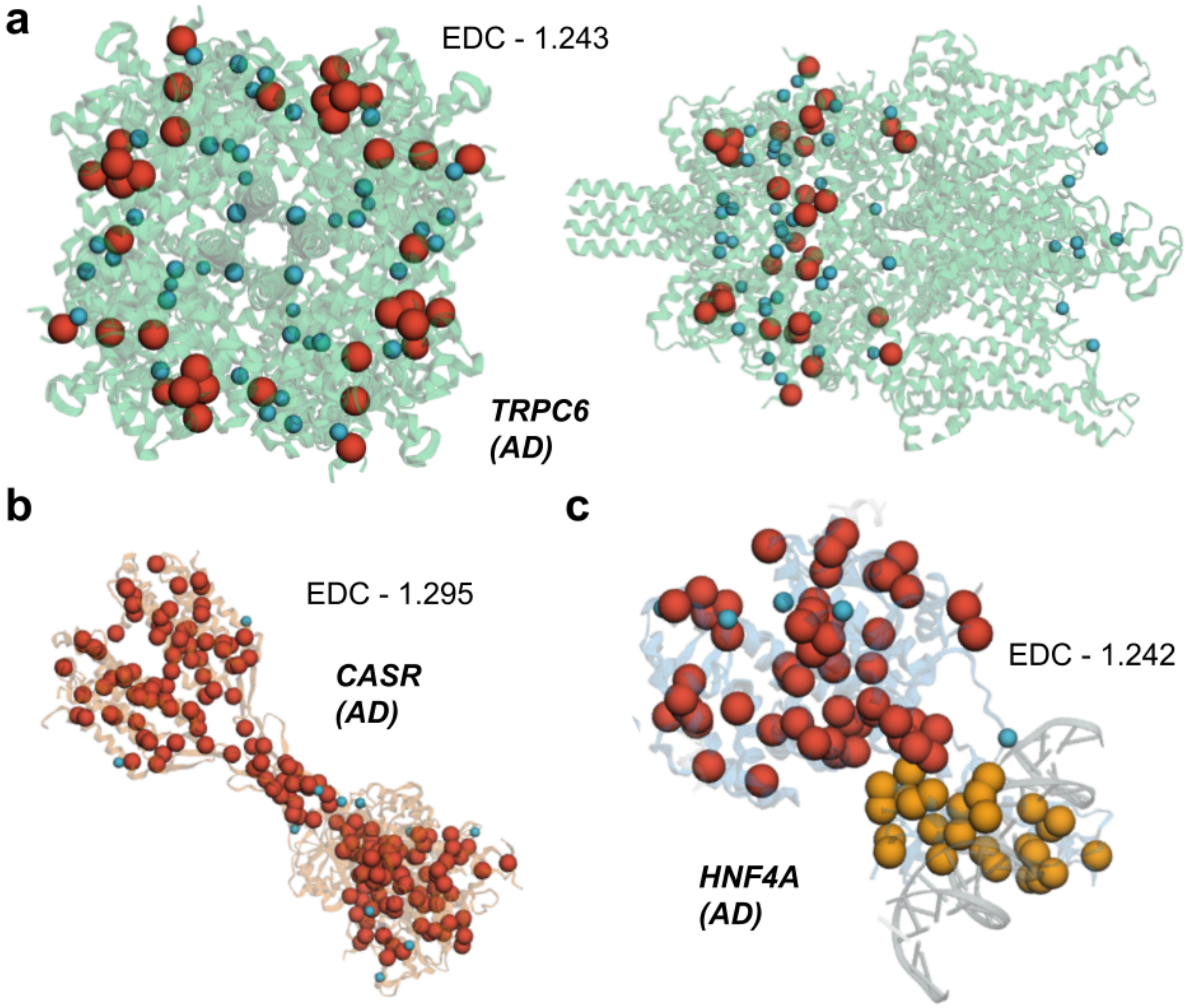

Figure S8. Pillai and Sayer., (2026).

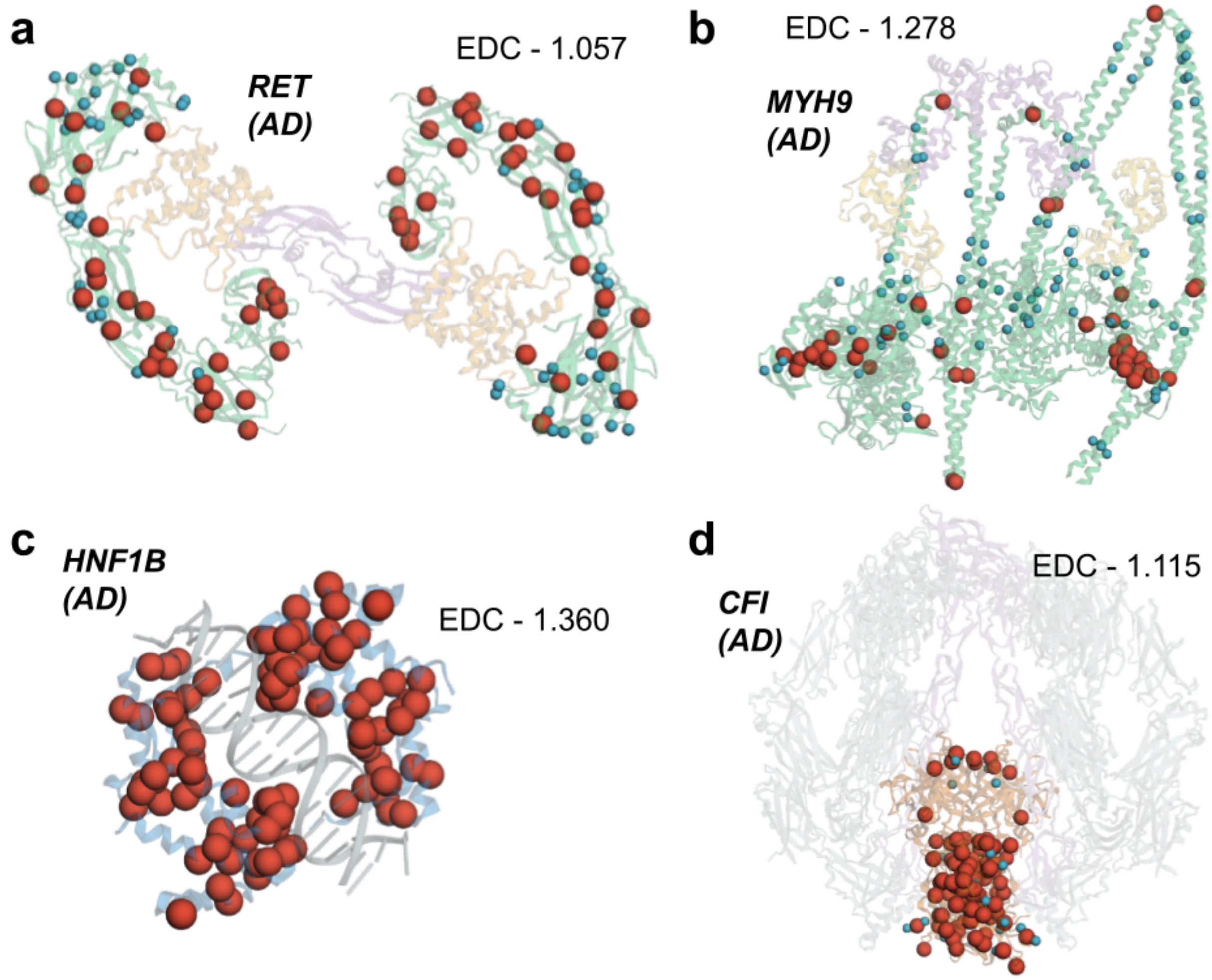

Figure S9. Pillai and Sayer., (2026).

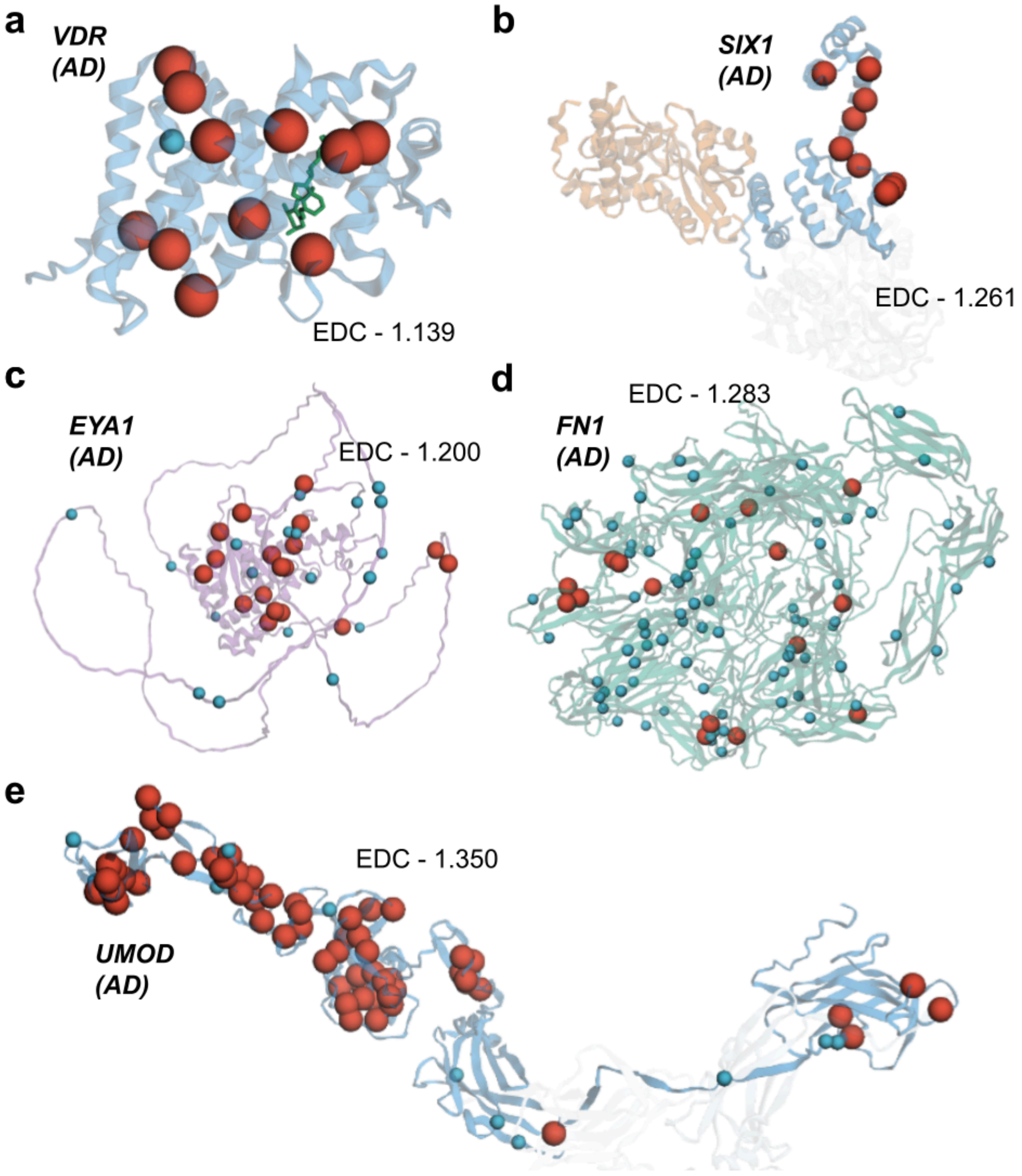
